# Gray Matter Volumes of the Superior Temporal Gyrus Link Preterm Birth and Developmentally Disordered Eye Gazing Patterns in Toddlers at Eighteen Months

**DOI:** 10.1101/2025.06.18.25329884

**Authors:** Yanan Su, Guangfei Li, Shanmei Wang, Dongmei Hao, Clara S. Li, Yiyao Ye-Lin, Xiaolin Wang, Ruolin Zhang, Lin Yang, Chiang-Shan R. Li

**Author notes:** Address correspondence to: Dr. Guangfei Li, 220 Life Sciences Building, Beijing University of Technology, 100 Pingleyuan, Beijing 100124, China.

## Abstract

**Background:** Preterm birth involves structural brain changes and increases the risk of neurodevelopmental disorders, including social cognitive dysfunction as implicated in autism spectrum disorder (ASD). However, it remains unclear whether or how volumetric brain changes may impact the risk of social cognitive dysfunction in toddlers of preterm birth.

**Methods:** We curated data of 569 toddlers approximately 18 months of age, including 76 with preterm (PB) and 493 with term (TB) birth, from the developing Human Connectome Project. We processed the imaging data, collected at birth, and investigated group differences in gray matter volume (GMV) of the brain and eye-tracking data collected at 18 months as well as the interrelationships amongst birth age, GMVs, and eye-tracking markers of ASD.

**Results:** In a covariance analysis with age at scan, total intracranial volume, sex, and number of embryos at gestation as covariates, PB demonstrated higher GMV in bilateral superior temporal gyri (STG). Right STG GMV’s were negatively correlated with birth age and positively with the proportion of looking at faces and mouths in PB, but not in TB. Further, path analyses suggested right STG GMV at birth as a marker of preferential face and mouth viewing in PB at 18 months.

**Conclusions:** The findings associate earlier birth age with atypical volumetrics of the right STG and eye gazing patterns in preterm children at 18 months. Longitudinal studies are needed to examine whether these neural and behavioral markers may reflect risks of social cognitive dysfunction in children with neurodevelopmental disorders, including ASD.

## **1.** Introduction

### 1.1. Preterm birth, brain insults, and developmental disorders

Preterm birth is a leading causes of infant mortality and morbidity, including adverse developmental outcomes, especially for those born with a very short gestational age (Platt, 2014). These include cerebral palsy, epilepsy, developmental coordination disorders, cognitive deficits, as well as socio-emotional and learning difficulties in childhood (Johnson et al., 2015; Moreira et al., 2014; Orchinik et al., 2011; Saigal and Doyle, 2008). Mid and late gestational stages are critical to fetal development, with brain growth increasing exponentially through the second half of pregnancy (Andescavage et al., 2017; Hinton et al., 2024; Li et al., 2025; Yang et al., 2024). Structural and microstructural brain changes (Gonzalez-Perez et al., 2024; Inder et al., 2005) and the associated functional abnormalities (Disselhoff et al., 2024; França et al., 2024; Rogers et al., 2018) resulting from preterm birth are likely key causal factors of behavioral and psychiatric problems that develop through adulthood (Johnson and Marlow, 2011).

Neurodevelopmental disorders and cognitive deficits are frequent outcomes of preterm birth (Bhutta et al., 2002). Preterm relative to full-term infants have two-fold the risk of neurodevelopmental disorders at age 2 years(Johnson et al., 2015). Preterm birth has been associated with risks of depression (Nomura et al., 2007; Patton et al., 2004), attention deficit or hyperactivity disorder (Linnet et al., 2006), and autism spectrum disorders (ASD) (Bokobza et al., 2019; Crump et al., 2021; D’Onofrio et al., 2013; Harel-Gadassi et al., 2018; Johnson et al., 2010; Johnson and Marlow, 2011; Khachadourian et al., 2023). The risks are inversely proportional to gestational age (Marlow et al., 2005; Marret et al., 2013; Schendel and Bhasin, 2008), with children of extremely (gestational age < 28 weeks) and very (28 to 32 weeks) preterm birth (Delobel-Ayoub et al., 2009) particularly vulnerable. For instance, earlier studies reported a nearly 30% higher risk of ASD in preterm children (Wang et al., 2017a) and four times the risk in those born at less than 27 weeks (Kuzniewicz et al., 2014), as compared to full-term children.

### 1.2. Behavioral markers of neurodevelopmental disorders

Early behavioral markers of neurodevelopmental disorders (NDD) include delays or atypicalities in sensory processing, motor skills, language development, social engagement, as well as repetitive behaviors or difficulties with attention and object use (Micai et al., 2020). Although no behavioral makers are diagnostic of a specific disorder, the hypothesis of social brain dysfunction has drawn significant attention in the research of the etiologies of ASD (Flores et al., 2025; Sato et al., 2023). Notably, the manifestation of social brain dysfunction may depend on age, and it does not rule out the possibility of brain-wide pathology and broader behavioral disarray in ASD (Gliga et al., 2014). Further, social cognitive dysfunction is observed in NDD other than ASD, including epilepsy, attention deficit hyperactivity and specific learning disorders (Novak et al., 2024; Pastorino et al., 2021). Indeed, studies have suggested shared and converging molecular and functional pathways for many NDD (Parenti et al., 2020). Nonetheless, social brain dysfunction has most been investigated for ASD, and we focused on studies of a critical behavioral marker – eye movement during exposure to social scenes – of social brain dysfunction to highlight the scientific premise of the current work.

As with many other NDD, ASD is characterized by persistent social and communication difficulties (First, 2013), with age-different symptoms, for instance, lack of eye contact and gaze-following in children (Reynolds and Culican, 2023; Stuart et al., 2023) and of social interaction in adolescents and adults (Au et al., 2021; Corbett et al., 2014; Simon et al., 2021). Abnormal gaze patterns suggest a fundamental problem with attention to socially relevant versus irrelevant information (Frazier et al., 2017). An eye-tracking study showed that the percentage of time fixated on the eyes was significantly decreased in preschool children with ASD, relative to controls (Kong et al., 2022). Infants and children later diagnosed with ASD looked less at faces and showed less eye contact and gaze-following behavior (Stuart et al., 2023). Many other studies reported that young children with ASD look significantly less at people’s eyes and more at people’s mouths (Jones et al., 2008b; Tanaka and Sung, 2016; Zwaigenbaum et al., 2005). Adolescents and adults with ASD had similar gaze patterns in the face recognition task, including reduced attention to the face and to the eyes (Mastergeorge et al., 2021; Setien-Ramos et al., 2023). Together, these findings suggest altered eye gaze patterns during exposure to human faces and social scenes and the utility of eye tracking in revealing social cognitive dysfunction in NDD, including ASD.

### 1.3. Sex differences in NDD

NDD show sex differences in clinical presentations (Nowak and Jacquemont, 2020) and underlying biological mechanisms (Breach and Lenz, 2023; Dan, 2021; May et al., 2019). NDD occur more often in boys than girls (Nowak and Jacquemont, 2020); for instance, males as compared to females are more likely to be diagnosed with ASD (Elsabbagh et al., 2012; Halladay et al., 2015), with a ratio of approximately 3:1 (Loomes et al., 2017) and boys also diagnosed at an earlier age than girls (Giarelli et al., 2010; Rutherford et al., 2016). Other studies documented sex differences in the clinical manifestations of ASD (Eckerd, 2020; Evans et al., 2019; Hull et al., 2017; Hull et al., 2020; Lawson et al., 2018; Ros-Demarize et al., 2020).

Eye-tracking studies have also reported sex differences in the behavioral markers of NDD. For instance, an experimental study of children 8 years of age showed that, though both deficient in social attention relative to neurotypical counterparts, females appeared to be less impaired than males with ASD (Harrop et al., 2019). Males but not females around 19 years old with high autistic-like traits showed worse eye gaze patterns than those with lower traits (Whyte and Scherf, 2018). Male infants with risk of ASD paid more attention to the mouth, whereas females with the risk paid little attention to the mouth but more to the eyes, as compared to their control counterparts (Kleberg et al., 2019), when viewing human faces. Thus, it would seem important to investigate sex differences in these behavioral markers of NDD.

### 1.4. Gray matter volumetric markers of ASD

Magnetic resonance imaging has been widely used to study brain volumetric markers of NDD, including ASD (Liloia et al., 2024). Understanding brain development in NDD requires characterization of the changes in gray matter volume (GMV) across ages. A meta-analysis of more than 100 MRI studies showed that GMVs increase significantly from the second trimester, peak in growth rate at about five months, and start to decline at about 6 years of age (Bethlehem et al., 2022). Many studies have reported atypical GMV in individuals with ASD, with regional changes related to the core symptoms (Xu et al., 2024). For instance, children/adolescents with ASD were more likely than adults to have higher GMV in bilateral fusiform gyrus, right cingulate gyrus, and insula, relative to their controls (Duerden et al., 2012). Adults with ASD have higher gray matter density in the precentral gyrus (Winter, 2016) and lower GMV in the left parahippocampal gyrus (Yaxu et al., 2020). Children with ASD showed excessive brain volume growth as compared with typically developing children (Courchesne et al., 2007; de Jong et al., 2021; Hazlett et al., 2011), especially in the temporal and frontal lobes (Mostofsky et al., 2007; Radua et al., 2011; Wang et al., 2017b). This overgrowth is not present at birth but appears late in the first year of life (Hazlett et al., 2005; Shen et al., 2013). Thus, whereas the findings of regional structural abnormalities may vary with age, enlarged GMV appeared to common feature in ASD (Wang et al., 2022).

### 1.5. The present study

Preterm birth may cause behavioral and structural brain changes that reflect the risk of NDD, including ASD. Here, we examined regional volumetric differences in preterm as compared to full-term infants at term-equivalent age, and eye gaze patterns during exposure to social scenes, a maker of social cognitive dysfunction, at 18 months of age, as well as sex differences in the abnormalities. We also explored whether the volumetric abnormalities support the impact of pre-term birth on eye gaze indicators of ASD risk. We broadly hypothesized volumetric differences in the social brain circuits, including the superior temporal cortex, in preterm vs. full-term infants and sex differences in how these volumetric abnormalities relate eye gaze behaviors.

## 2. Methods

### 2.1. Participants and assessments

We employed data collected from the Developing Human Connectome Project (dHCP, www.developingconnectome.org/). This project has received ethical approval (14/LO/1169, IRAS 138070), with written informed consent obtained from the parents.

The dHCP included a total of 887 MR images from 783 subjects, with 101 subjects scanned twice, once at 26.7 to 42.7 (33.8 ± 2.8) weeks and a second time at 29.3 to 44.9 (41.1 ± 2.1) weeks, five subjects scanned three times, each at 28.6 to 34.9 (31.9 ± 3.2) weeks, 29.3 to 35.6 (32.4 ± 4.4) weeks, and 39.9 to 42.7 (41.7 ± 1.2) weeks, and one subject scanned four times, first at 33.3 weeks and last at 42.3 weeks. We excluded 1) those without a radiology score or with a score > 2, indicating gross structural abnormality, punctate lesions or other significant radiological findings (n = 218), 2) those scanned using sedation (n = 5), 3) those scanned before 37 weeks of postmenstrual age (PMA, i.e., gestational age + chronological age, n = 91), and 4) those scanned twice (n=4; the one with scan time closer to the 38 weeks of PMA was included in the analyses). Thus, a total of 569 subjects were included in this study and categorized according to birth age: < 37 weeks as the preterm birth (PB, n = 76) and ≥ 37 weeks as term birth (TB, n = 493) group. We also distinguished males and females with four subgroups: PB males (n = 46, PBM), PB females (n = 30, PBF), TB males (n = 264, TBM), and TB females (n = 229, TBF).

The Quantitative Checklist for ASD in Toddlers (Q-CHAT), a 25-item screening questionnaire, was used to assess autistic traits (Allison et al., 2008). A higher score on the Q-CHAT indicates more behaviors and traits in line with those of ASD. However, not all of the participants had Q-CHAT scores. Of the entire cohort, 34 PBM, 199 TBM, 21 PB, 195 TBF were evaluated with the Q-CHAT (**Table 1**, lower panel). Demographic and clinical information of the four groups is summarized in **Table 1**. As expected, PB relative to TB showed lower birth age, smaller TIV, higher number cases of non-singlet birth and with a radiological score of 2 rather than 1.

**Table 1.** Demographic and clinical measures of the participants.

| Characteristic | PBM<br>(n=46) | TBM<br>(n=264) | PBF<br>(n=30) | TBF<br>(n=229) | Two-way ANOVA* |  |  |  |  |  |
| --- | --- | --- | --- | --- | --- | --- | --- | --- | --- | --- |
|  |  |  |  |  | Group |  | Sex |  | Interaction |  |
|  |  |  |  |  | F | p | F | p | F | p |
| birth_age | 32.8±3.4 | 40.0±1.2 | 31.4±3.2 | 40.1±1.3 | 1417 | 0.000 | 10.17 | 0.002 | 12.19 | 0.001 |
| scan_age | 40.7±1.9 | 41.3±1.7 | 41.4±1.9 | 41.6±1.8 | 2.90 | 0.089 | 4.60 | 0.032 | 0.53 | 0.465 |
| TIV | 385.7±9.3 | 392.6±6.7 | 385.7±8.2 | 391.5±7.3 | 49.26 | 0.000 | 0.33 | 0.564 | 0.35 | 0.553 |
| Singlet (Y/N) | 29/17 | 254/10 | 20/10 | 219/10 | 94.34 | 0.000 | 0.23 | 0.634 | 0.43 | 0.512 |
| RS: 1/2 | 26/20 | 156/108 | 13/17 | 160/69 | 5.79 | 0.016 | 0.04 | 0.842 | 3.93 | 0.048 |

| Characteristic | PBM<br>(n=34) | TBM<br>(n=199) | PBF<br>(n=21) | TBF<br>(n=195) | Two-way ANOVA* |  |  |  |  |  |
| --- | --- | --- | --- | --- | --- | --- | --- | --- | --- | --- |
|  |  |  |  |  | Group |  | Sex |  | Interaction |  |
|  |  |  |  |  | F | p | F | p | F | p |
| EyeTrAge | 21.5±3.3 | 19.0±2.2 | 21.0±1.6 | 19.0±1.7 | 29.96 | 0.000 | 0.52 | 0.472 | 0.38 | 0.538 |
| Mo_edu_age | 21.9±2.9 | 23.3±4.2 | 23.4±3.5 | 23.6±4.6 | 0.00 | 0.986 | 1.00 | 0.317 | 0.86 | 0.355 |
| Fa_edu_age | 20.8±3.0 | 23.2±4.3 | 23.6±2.6 | 23.8±5.3 | 3.34 | 0.068 | 6.84 | 0.009 | 2.34 | 0.127 |
| Q-CHAT | 31.2±12.9 | 31.4±9.1 | 29.4±8.9 | 29.2±7.6 | 0.39 | 0.530 | 1.94 | 0.164 | 0.22 | 0.883 |

Note: These measures are separated in four groups, according to their sample sizes.
PBM: male in PB; TBM: male in TB; PBF: female in PB; TBF: female in TB; birth\_age: gestational age at birth, in weeks; scan\_age: postmenstrual age at scan, in weeks;
TIV: total intracranial volume (mm<sup>3</sup>); Singlet (Y/N): Yes/No: singleton/multiple; RS: radiology score – only those with an RS of 1 or 2 were included in the study; EyeTrAge: age at eye-tracking experiment, in months; Mo/Fa\_edu\_age: age when mother/father was last in continuous full-time education; QCHAT: total Q-CHAT score. \*Two-way ANOVA with age at scan and number of embryos at gestation as covariates.

### 2.2 Image data acquisition, preprocessing, and analyses

Images were acquired on a 3T Phillips 3-Tesla Achieva system in a natural sleep state, using a dedicated neonatal brain imaging system, including a neonatal 32-channel receive coil and a customized setup (Hughes et al., 2017).

All images were acquired in Evelina Newborn Imaging Centre, Evelina London Children’s Hospital. T2-weight images were obtained using a TSE sequence with parameters of TR = 12000ms, TE = 156ms, SENSE factor 2.11 (axial) and 2.58 (sagittal), and resolution (mm) of 0.8 × 0.8 × 1.6. Note that T2 rather than T1 images were acquired because of their better contrasts of brain tissues in fetal or newborn brain (Makropoulos et al., 2018a).

All anatomical images were reviewed by an expert perinatal neuroradiologist, and scores of 1 and 2 were considered normal or having lesions unimportant for the analysis, and scores of 3 to 5 or Q indicated that the anatomical data were of poor quality and/or had lesions that would impact the analysis.

Bias-corrected T2-weight images were selected which were processed through the dHCP minimal preprocessing pipeline for Neonatal Cortical Surface Reconstruction (Makropoulos et al., 2018b). Additional preprocessing procedures for the images included extracting the brain using BET in the FMRIB software library (FSL) (Jenkinson et al., 2012), strictly recording the brain volume into the standard space of the CRL 38-week template (Gholipour et al., 2023) by ANTs (Avants et al., 2008), followed by segmentation using the atlas (Serag et al., 2012a; Serag et al., 2012b) used in Draw-EM (Makropoulos et al., 2014; Makropoulos et al., 2018b), and smoothing the gray matter, white matter, and cerebrospinal fluid obtained from segmentation to compute the volume for statistical analysis. TIV was calculated as the sum of the volumes of gray matter, white matter, and cerebrospinal fluid.

For the entire cohort of 569 subjects (PBM=46, TBM=264, PBF=30, TBF=229), we performed a two-sample t test of PB vs. TB, PBM vs. TBM, and PBF vs. TBF with age at scan, TIV, and number of embryos at gestation as covariates. We also performed a group X sex ANOVA with the same covariates to examine the interaction term for sex differences. For all analyses, we assessed the results at voxel p < 0.001, uncorrected in combination with a cluster p < 0.05 FWE-corrected, based on Gaussian random field theory as implemented in SPM.

### 2.3 Eye-Tracking data acquisition and analyses

Spatial characteristics of gaze were collected from a total of 609 subjects (78% of the sample) at a median age of 18 months and 12 days (range 17 months, 8 days to 34 months, 15 days) during the “50 faces” task, derived from the “50 People, One Question” project (Krolak-Salmon, 2011), where the participants watched a 41-second video of a street interview. Eye-Tracking data were acquired using a Tobii TX-300 (Tobii AB, Sweden) gaze tracking system, with the subject sitting approximately 60 cm from the screen. The dHCP employed a customized stimulus series of animated video clips to evaluate visual attention. The video covered primarily the interviewee’s face and body parts above the chest and the framing focused on the interviewee. The figures in the background were relatively ambiguous with non-social stimulation (Braithwaite et al., 2023). The sound of the original video was removed and replaced by classical music to remove the potential influences of language (**Figure 1b**).

**Figure 1.**
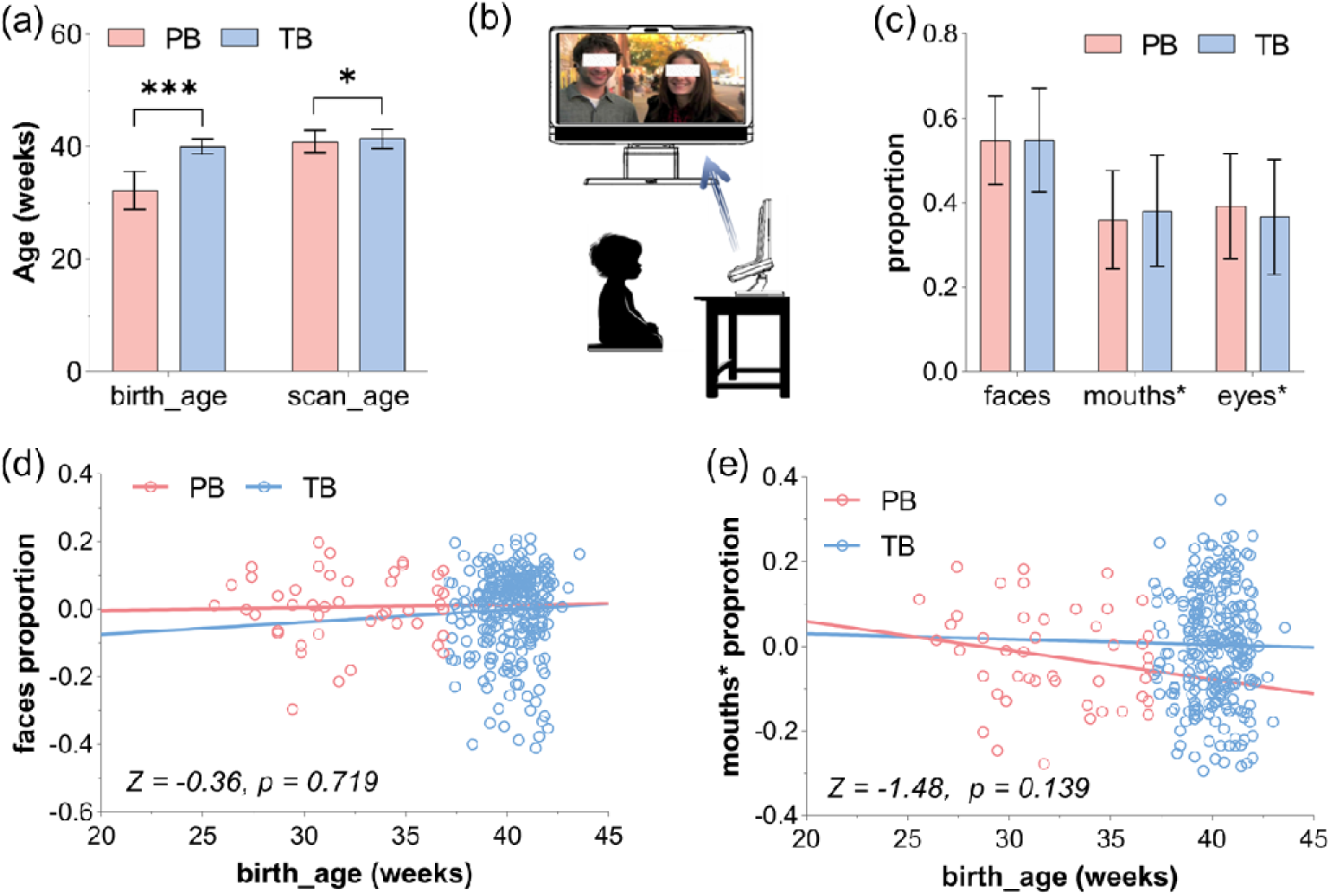
(a) Histogram of age of birth and age at scan in PB and TB: mean ± SD. *p<0.05; ***p<0.001. **(b)** Eye-tracking experiment of the “50 faces” task. **(c)** Histogram of the proportion of looking at faces, mouths* (faces - eyes), and eyes* (faces - mouths) in PB and TB: mean ± SD. **(d)** Regression of age at birth vs. proportion of looking at faces in PB vs. TB. **(e)** Regressions of age at birth vs. proportion of looking at mouths* in PB vs. TB. Y coordinates represent the residuals after regressing out age at scan, sex, and number of embryos at gestation.

Participants’ eye positions were tracked throughout the viewing period, and eye-tracking data were provided by the dHCP (Braithwaite et al., 2023). Specifically, areas-of-interest (AOIs) were manually drawn frame by frame around face, eyes, mouth, people in the background, using Motion software (Apple Inc, USA), with the key variables of mean proportion, duration, and peak duration of looking at each AOI recorded (Braithwaite et al., 2023). Face AOI included eyes and mouth. Here, we focused on proportion of gaze time spent on faces as well as proportion of “faces minus eyes” and “faces minus mouths” for evaluation of eye-tracking performance and for correlation with neural and clinical metrics (**Table 1**). We considered “faces – eyes” and “faces – mouths” each as an indicator of mouths- and eyes- gazing because participants may demonstrate transitional fixations on their gaze shift to the mouths/eyes; thus, we included these transitional fixations as part of their effort in looking at mouths and eyes, respectively. We used faces, eyes* (faces – mouths), and mouths* (faces – eyes) proportion in evaluating the group differences in eye gazing metrics and the correlation of these metrics with clinical and neural measures. We also reported AccuracyDeg or PrecisionDeg, each defined as the spatial displacement of recorded gaze from the point fixated and variability in consecutive samples on the same fixation point, respectively, which reflected the general quality of eye tracking data (Braithwaite et al., 2023). **Supplementary Table S1** show all other recorded eye-tracking measures.

For the subgroup of toddlers evaluated with the eye-tracking task (PBM=28, TBM=128, PBF=17, TBF=138), we performed correlation analysis of eye-tracking metrics and the GMVs of brain regions identified of whole-brain group analyses, with age at scan, TIV, sex, and number of embryos at the time of gestation as covariates. We evaluated the results of correlation with a p value corrected for the number of tests in a specific set of analysis.

### 2.4 Path analyses

We employed path analysis to evaluate how severity of preterm birth, eye-tracking metrics and the neural correlates were inter-related (Chen et al., 2008; Hu and Bentler, 1995; Ji et al., 2024; Li et al., 2024; Li et al., 2021a; Li et al., 2021b; Li et al., 2023). Specifically, we tested whether the volumetric markers may inter-link birth age and eye-tracking measures of significance (see “Results”) in the subgroup of toddlers evaluated with the eye-tracking task (PBM=28, TBM=128, PBF=17, TBF=138). Model fit was assessed with standard fit indices which included the Root Mean Square Estimation of Approximation (RMSEA, < 0.08 for an acceptable fit), Chi-square (χ^2^/df, < 3), Comparative Fit Index (CFI, > 0.9), and Standardized Root Mean Square Residual (SRMR, < 0.06). We considered all possible path models for the sake of completeness, although, with imaging and eye-tracking data each collected at birth and at around 18 months of age, only models birth age ➔ volumetric markers ➔ eye-tracking metrics are conceptually valid.

## 3. Results

### 3.1. PB vs. TB in clinical and eye-tracking metrics

Clinical characteristics as well as the statistics of group x sex ANOVA are summarized in **Table 1**, with a narrative description provided in the **Supplementary Results**. Here, we described on the main findings.

The Q-CHAT score showed no significant group (F = 0.39, p = 0.530) or sex (F = 1.94, p = 0.164) main effect, nor interaction effect (F = 0.22, p = 0.883).

For eye-tracking metrics, AccuracyDeg or PrecisionDeg showed no significant group (all p’s > 0.182) or sex (all p’s > 0.672) main effect or group by sex interaction effect (all p’s > 0.559), suggesting no significant differences in the quality of eye-tracking data. Among the three primary metrics – proportion of faces, eyes*, and mouths*, none showed a significant group or sex main or group by sex interaction effect at a corrected threshold of p < 0.05/3 = 0.017. At an uncorrected threshold, only the proportion of looking at eyes* showed a significant group main effect (F = 3.90, p = 0.049, uncorrected), but not sex main (F = 0.00, p = 0.976) or interaction (F = 0.55, p = 0.460) effect (**Table 2**). Specifically, PB relative to TB showed a higher proportion of looking at eyes* (0.39 ± 0.12 vs. 0.37 ± 0.14; F = 3.90, p = 0.049).

**Table 2.** Eye-tracking measures of the participants.

| Characteristic | PBM<br>(n=28) | TBM<br>(n=128) | PBF<br>(n=17) | TBF<br>(n=138) | Two-way ANOVA* |  |  |  |  |  |
| --- | --- | --- | --- | --- | --- | --- | --- | --- | --- | --- |
|  |  |  |  |  | Group |  | Sex |  | Interaction |  |
|  |  |  |  |  | *F* | *p* | *F* | *p* | *F* | *p* |
| faces_Prop | 0.54±0.12 | 0.54±0.12 | 0.55±0.09 | 0.55±0.13 | 0.19 | 0.667 | 0.10 | 0.748 | 0.02 | 0.883 |
| mouths*_Prop | 0.36±0.12 | 0.38±0.13 | 0.36±0.12 | 0.38±0.13 | 2.26 | 0.133 | 0.16 | 0.685 | 0.03 | 0.857 |
| eyes*_Prop | 0.39±0.13 | 0.36±0.13 | 0.39±0.12 | 0.37±0.14 | 3.90 | 0.049 | 0.00 | 0.976 | 0.55 | 0.460 |
| AccuracyDeg | 1.88±0.82 | 1.69±0.76 | 1.81±1.59 | 1.68±0.63 | 1.79 | 0.182 | 0.18 | 0.672 | 0.09 | 0.767 |
| PrecisionDeg | 1.54±0.47 | 1.49±0.41 | 1.59±0.42 | 1.48±0.40 | 1.28 | 0.259 | 0.07 | 0.789 | 0.34 | 0.559 |

### **3.2** Correlation amongst age at birth, Q-CHAT score, and eye-tracking metrics

As shown in **Supplementary Table S2** (and **Supplementary Figure S1** in matrix format), age at birth was not significantly correlated with Q-CHAT score in any group at a corrected threshold (all p’s > 0.044). None of the three eye-tracking measures showed a significant correlation with Q-CHAT score in any group (all p’s > 0.128). The statistics of correlation of Q-CHAT score with other eye-tracking metrics were summarized in the **Supplementary Table S3.**

In the correlation of birth age and eye-tracking metrics, the proportion of looking at mouths* was significantly and negatively correlated with age at birth in PBF (r = -0.67, p = 0.009) but not in PBM (r = 0.12, p = 0.567), at a relaxed threshold of p < 0.05/3 = 0.017. The slope test showed a significant sex difference (PBF vs. PBM) in the slope of the linear regressions (Z = -2.78, p = 0.005). None of the three primary eye-tracking measures showed a significant correlation with age at birth in any other group (all p’s > 0.071). The statistics of correlation of age at birth with other eye-tracking metrics were summarized in the **Supplementary Table S4.**

### 3.3 Gray matter volumes

For the entire cohort of 569 subjects (PBM=46, TBM=264, PBF=30, TBF=229), we performed a two-sample t test for the whole brain of PB vs. TB, PBM vs. TBM, and PBF vs. TBF with age at scan, TIV, and number of embryos at gestation as covariates. Evaluated at voxel p < 0.001, uncorrected in combination with a cluster p < 0.05 FWE-corrected, the findings are shown for all PB vs. TB in **Figure 2a**, for PBM vs. TBM in **Figure 2b**, and for PBF vs. TBF in **Figure 2c**. The clusters are summarized in **Table 3**. At the same threshold, we did not observe any clusters showing significant interaction [i.e., (PBF – TBF) – (PBM – TBM)] or sex main effect in a group by sex ANOVA for the whole brain.

**Figure 2.**
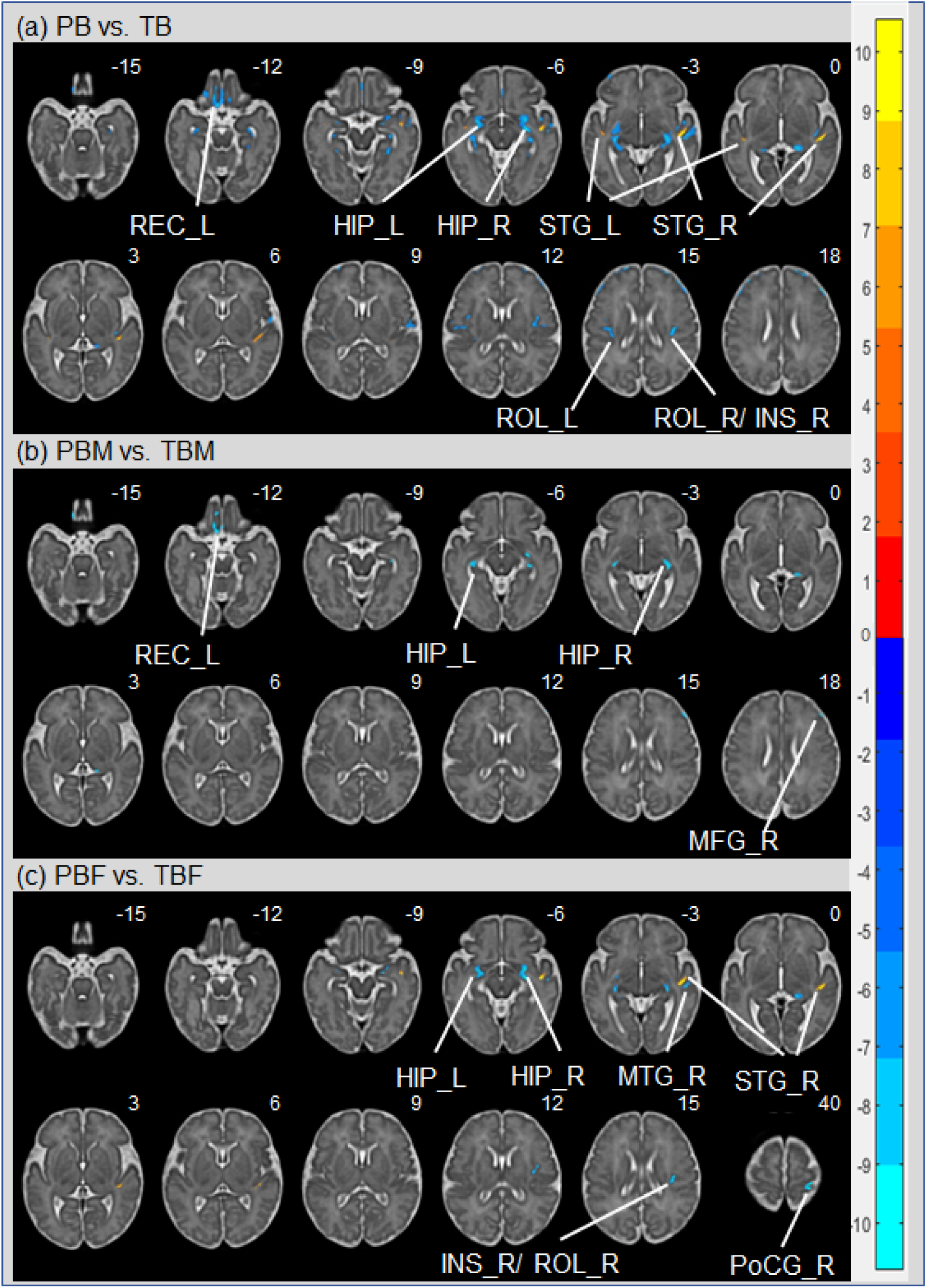
Regional GMV’s with group differences. **(a)** PB vs. TB. Color bars show voxel T values: warm: PB > TB, cool: PB < TB. **(b)** PBM vs. TBM; warm: PBM > TBM, cool: PBM < TBM. **(c)** PBF vs. TBF; warm: PBF > TBF, cool: PBF < TBF. All comparisons were with age at scan, TIV, and number of embryos at gestation as covariates, evaluated at voxel p < 0.001, uncorrected, cluster p < 0.05 FWE-corrected. Clusters are overlaid on the T2 structural image in neurological orientation: right = right. L: left; R: right. STG: superior temporal gyrus; HIP: Hippocampus; REC: Gyrus rectus; ROL: Rolandic operculum; INS: Insula. MFG: middle frontal gyrus; PoCG: Postcentral gyrus; MTG: Middle temporal gyrus.

**Table 3.** GMV differences in PB vs. TB, PBM vs. TBM, and PBF vs. TBF.

| Region | Cluster size | Peak Level (T) | Cluster FWE P- value | MNI coordinates (mm) |  |  |
| --- | --- | --- | --- | --- | --- | --- |
|  |  |  |  | x | y | z |
| *PB > TB* |  |  |  |  |  |  |
| STG_R | 543 | 10.56 | 0.000 | 32 | 6 | -4 |
| STG_L | 105 | 7.38 | 0.000 | -28 | 6 | -4 |
| *PB < TB* |  |  |  |  |  |  |
| HIP_R | 1302 | 10.12 | 0.000 | 19 | 8 | -6 |
| HIP_L | 1151 | 8.20 | 0.000 | -18 | 10 | -5 |
| REC_L | 829 | 9.23 | 0.000 | -4 | 27 | -12 |
| ROL_R/ INS_R | 426 | 9.65 | 0.000 | 25 | 8 | 14 |
| ROL_L | 357 | 8.33 | 0.000 | -25 | 6 | 16 |
| *PBM < TBM* |  |  |  |  |  |  |
| REC_L | 266 | 7.32 | 0.000 | -6 | 42 | -14 |
| HIP_R | 259 | 7.45 | 0.000 | 20 | 1 | -4 |
| HIP_L | 161 | 7.15 | 0.000 | -22 | 1 | -5 |
| MFG_R | 85 | 7.70 | 0.000 | 32 | 32 | 21 |
| *PBF > TBF* |  |  |  |  |  |  |
| STG_R | 425 | 9.77 | 0.000 | 32 | 6 | -5 |
| *PBF < TBF* |  |  |  |  |  |  |
| HIP_R | 599 | 8.91 | 0.000 | 19 | 8 | -6 |
| INS_R/ ROL_R | 132 | 7.90 | 0.000 | 24 | 2 | 14 |
| PoCG_R | 100 | 8.03 | 0.000 | 21 | -6 | 40 |
| MTG_R | 100 | 7.35 | 0.000 | 34 | 3 | -4 |
| HIP_L | 96 | 6.60 | 0.000 | -14 | -6 | -2 |

Note: Brain regions were identified by reference to the atlas of (Gholipour et al., 2017). L: left; R: right; HIP: hippocampus; INS: Insula; MFG: Middle frontal gyrus; MTG: Middle temporal gyrus; PoCG: Postcentral gyrus; REC: gyrus rectus; ROL: Rolandic operculum; STG: superior temporal gyrus;

### 3.4 Correlation between age at birth, GMV’s, and eye-tracking metrics

#### 3.4.1 Regions showing higher GMV in PB vs. TB

PB relative to TB showed higher GMVs in bilateral superior temporal gyrus (STG). With age at scan, TIV, and number of embryos at gestation as covariates, age at birth was significantly and negatively correlated with right STG GMV for the entire cohort (r = -0.47, p < 0.001) and in PB (r = -0.37, p = 0.017) but not in TB alone (r = -0.03, p = 0.660), with the slope test confirming the difference in slope of the regression for PB vs. TB (Z = -2.18, p = 0.029) (**Figure 3a**). Age at birth was significantly and negatively correlated with left STG GMV only for the entire cohort (r = -0.33, p < 0.001) but not in PB (r = -0.16, p = 0.306) or in TB (r = -0.07, p = 0.257) alone.

**Figure 3.**
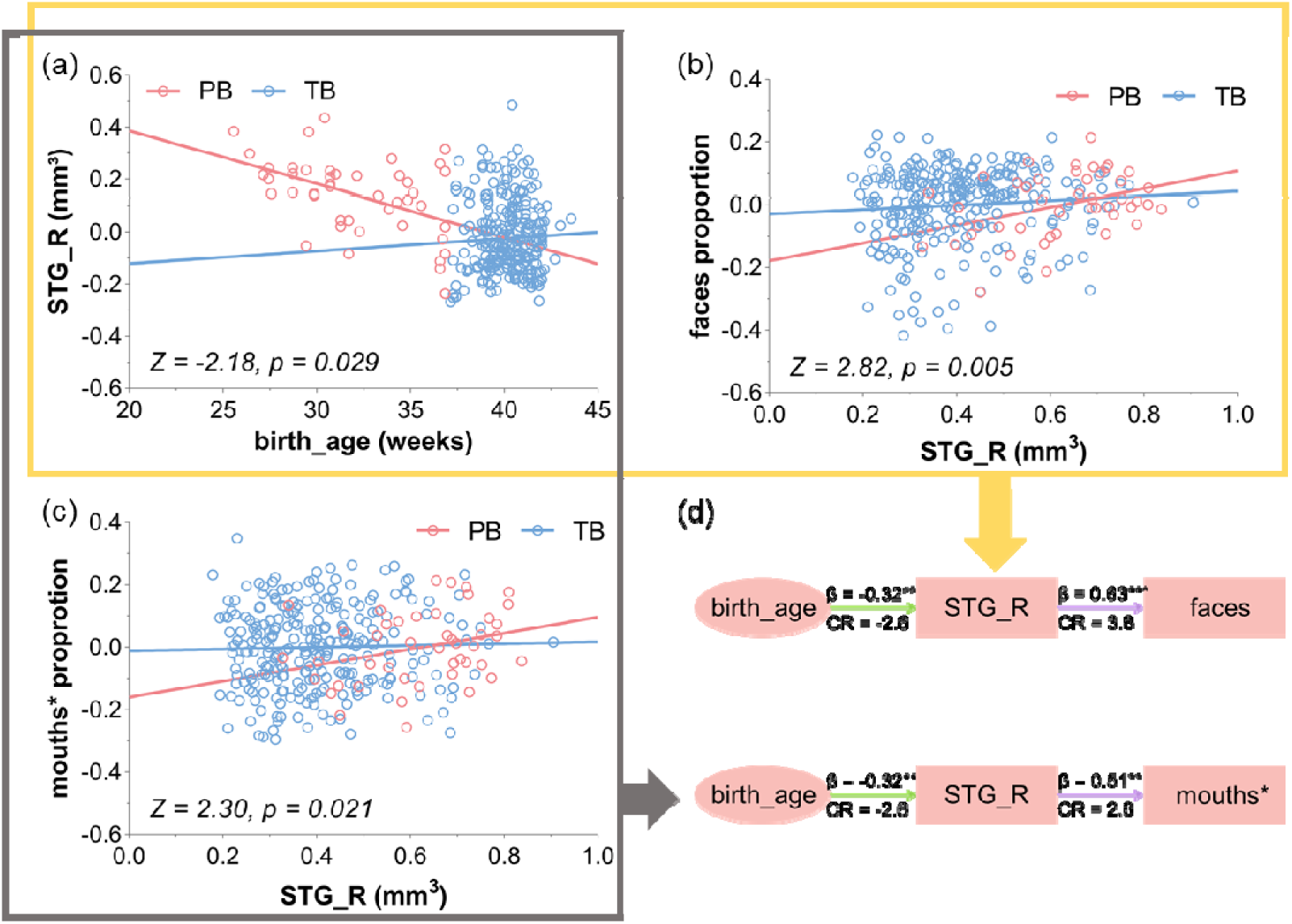
(a) Correlation of age at birth and STG_R GMV (as identified from whole-brain analysis; Figure 2a). **(b)** Correlation of STG_R GMV and proportion of looking at faces. **(c)** Correlation of STG_R GMV and proportion of looking at mouths*. **(d)** Two path analyses in PB of age at birth, STG_R GMV, and proportion of looking at faces or proportion of looking at mouths*. Purple and green arrow each indicates positive and negative effect. Y coordinates are the unstandardized residuals after regressing covariates: age at scan, sex, TIV and number of embryos at gestation. **p<0.01; ***p<0.001. CR: critical ratio = path coefficient/ standard error.

We next examined whether the right and left STG GMVs were related to eye-tracking metrics at a corrected p < 0.05/(2 regions x 3 metrics) = 0.008.

In the entire cohort, right STG GMV was not significantly correlated with any eye-tracking metric (all p’s > 0.037). Left STG GMV was significantly correlated with proportion of looking at faces (r = 0.18, p = 0.002) and eyes* (r = 0.22, p < 0.001), but not mouths* (r = 0.04, p = 0.529).

In PB, right STG GMV showed a significant and positive correlation with the proportion of looking at faces (r = 0.50, p = 0.001) but not mouths* (r = 0.39, p = 0.012) or eyes* (r = 0.18, p = 0.273). Left STG GMV showed a significant and positive correlation with proportion of looking at faces (r = 0.41, p = 0.007), but not with proportion of looking at mouths* or eyes* (all p’s > 0.094).

In TB, right STG GMV was not correlated with any of the three metrics (all p’s > 0.266). Left STG GMV was significantly and positively correlated with proportion of looking at eyes* (r = 0.20, p = 0.001), but not mouths* (r = 0.04, p = 0.474) or faces (r = 0.15, p = 0.014).

These correlations as well as slope tests of PB vs. TB are shown in **Supplementary Figure S2.** Notably, the regression of right STG GMV with faces and mouths* looking differed significantly in slope between PB and TB (**Figure 3b** and **3c**). The r and p values of all regressions are summarized in **Supplementary Table S5**.

#### 3.4.2 Regions showing higher GMV in TB vs. PB

TB relative to PB showed higher GMVs in bilateral hippocampus, left gyrus rectus, and bilateral rolandic operculum.

With the same covariates, age at birth was significantly and positively correlated with GMVs of bilateral hippocampus and bilateral rolandic operculum across all subjects (all p’s < 0.001) and in TB (all p’s < 0.001). These correlations were also all positive for PB, although the r values were smaller likely due to the small sample size of this subgroup.

In contrast, none of these regional GMVs were significantly correlated with the any of the three eye-tracking metrics at a corrected threshold.

The r and p values of these correlations are summarized in **Supplementary Table S6a to S6e.**

### 3.5 Mediation analyses of age at birth, GMV’s, and eye-tracking metrics

In PB alone, right STG GMV showed both negative correlation with age (**Figure 3a**) at birth and positive correlation with proportion of looking at faces and mouths* (**Figure 3b** and **3c**). Thus, we performed path analyses to examine the inter-relationship between age at birth, right STG GMV and proportion of looking at mouths*. For the sake of completeness, we evaluated all six models, although the models with eye-tracking metrics as independent variables were chronologically unlikely.

The results of path analyses showed the models age at birth → right STG GMV → proportion of looking at faces and the model age at birth → right STG GMV → proportion of looking at mouths* with a significant and the best fit (**Figure 3d**). Other models did not show any significant model fit (**Supplementary Table S7** and **S8**; **Supplementary Figure S3** and **S4**).

### 3.6 Sex differences: exploratory analyses

As shown in **Figure 4**, slope tests showed sex difference in the relationship between age at birth and right STG GMV, and that between age at birth and proportion of looking at mouths*, both predominated by PBF.

**Figure 4.**
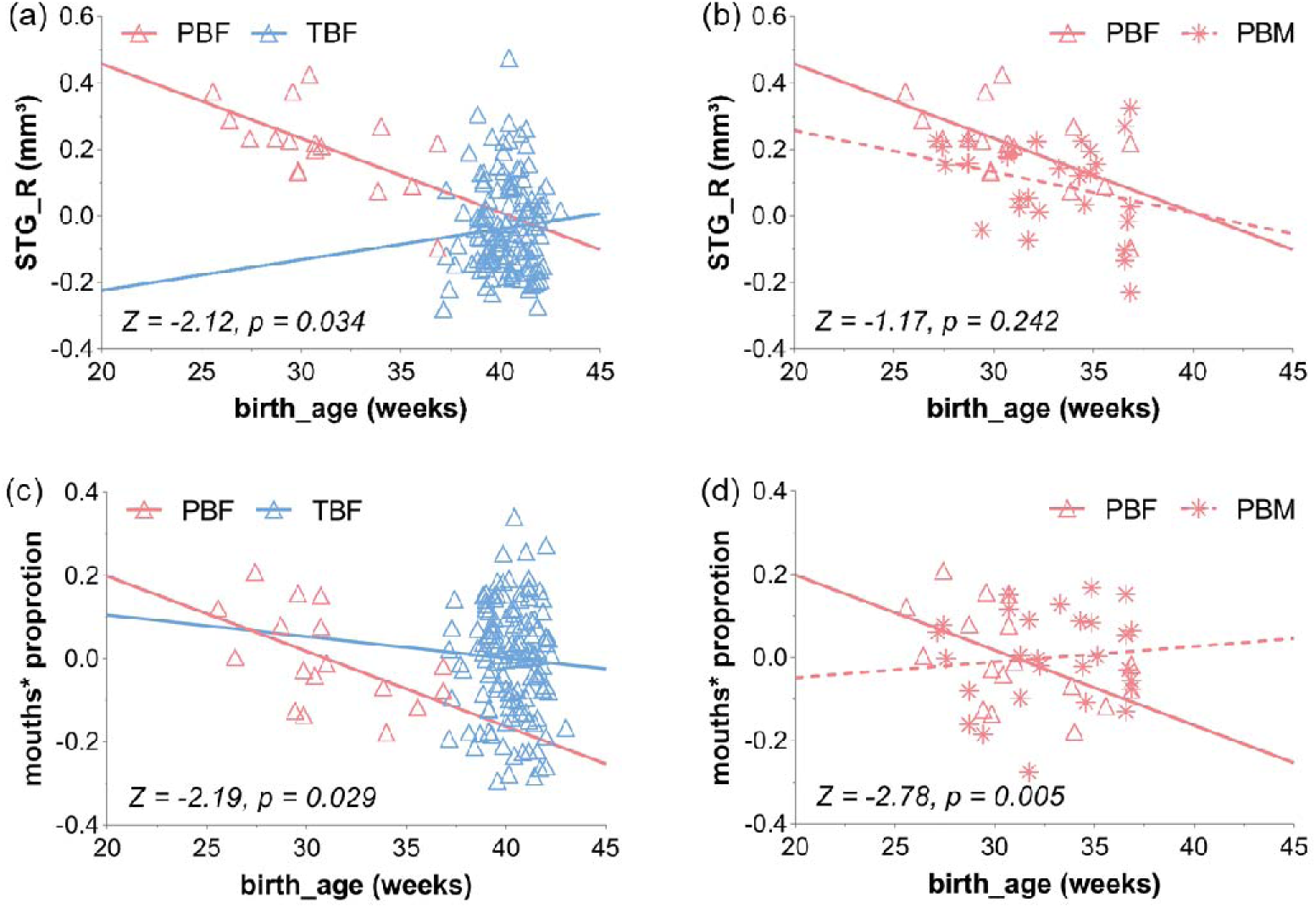
Sex differences in correlation of age at birth with STG_R GMV (as identified from whole-brain analysis; Figure 2a), and with proportion of looking at mouths*, using age at scan, TIV and the number of embryos at gestation as covariates. **(a)** PBF vs. TBF in correlation of age at birth with STG_R GMV. **(b)** PBF vs. PBM in correlation of age at birth with STG_R GMV. **(c)** PBF vs. TBF in correlation of age at birth with proportion of looking at mouths*. **(d)** PBF vs. PBM in correlation of age at birth with proportion of looking at mouths*. Y coordinates of **(a)** and **(b)** are the unstandardized residuals after regressing out: age at scan, TIV and number of embryos at gestation. Y coordinates of **(c)** and **(d)** are the unstandardized residuals after regressing two covariates: age at scan and number of embryos at gestation.

## 4. Discussion

In this study, we explored the brain volumetrics at birth and eye fixations while viewing a video of human interaction in PB and TB toddlers at 18 months of age. The results showed (1) larger GMV in right STG in PB vs. TB; (2) a positive correlation between the degree of prematurity and looking at faces and mouths* in PB; and (3) a path model with right STG GMV inter-linking birth age and preferential face- and mouth-looking in PB. PB relative to TB also showed smaller bilateral hippocampal GMVs but the volumetrics were not significantly associated with eye gazing measures. These results suggest that premature birth leads to atypical development of superior temporal cortex and gazing patterns that are indicative of social cognitive dysfunction and risks of NDD, including the ASD. We discussed the main findings in the below.

### 4.1 Preterm birth and higher STG GMV

Earlier birth age has been associated with larger brain volumes, likely reflecting accelerated brain growth in premature birth (Meredith Weiss et al., 2024). Here, preterm as compared to term newborns exhibited greater GMV in bilateral STG, consistent with an earlier report (Karolis et al., 2017) and studies suggesting vulnerability of temporal cortical and hippocampal regions to structural alterations in very preterm birth (Erdei et al., 2023; Ream and Lehwald, 2018). These structural markers of preterm birth may persist in adolescence and adulthood, with long-lasting effects on cognitive and affective function (Bjuland et al., 2014; Kelly et al., 2023). For instance, adolescents of preterm birth had higher cortical thickness in temporal gyrus (Ji et al., 2024; Nath et al., 2023). Adults of preterm birth had higher GMVs of the STG, prefrontal cortex, and occipital regions (Shang et al., 2019). In particular, structural changes in the posterior STG, which harbors the planum temporale or Wernicke area (Lindell and Hudry, 2013), the primary site of auditory and sensory speech processing (Hullett et al., 2016; Leff et al., 2009; Yi et al., 2019), may lead to deficits in speech and language development (Bhaya-Grossman and Chang, 2022; Eyler et al., 2012). Further, the STG is involved in processing social cues, including people’s face expressions, voices, gestures, and other contextual information (Adolphs, 2003; Paulus et al., 2005; Singer et al., 2004; Xu et al., 2009). Thus, insults to the STG as can occur during preterm birth may lead to social cognitive dysfunction, a hallmark of ASD and other NDD (Bigler et al., 2007; Pelphrey et al., 2004).

In the current sample, PB and TB did not differ in total Q-CHAT score, which we speculated may result from the low validity of Q-CHAT (Thabtah and Peebles, 2019) and the broader challenge of evaluating clinical features “diagnostic” of NDD in very young children, and/or the relatively small sample size of PB. With this caveat, we discussed briefly the literature of ASD. Relative to typically developing children, those with ASD showed higher temporal lobe GMV (Foster et al., 2015; Lukito et al., 2020; Piven et al., 1995; Retico et al., 2016; Wang et al., 2017b), with some studies implicating specifically the right temporal cortex across age groups (Carper and Courchesne, 2005; Carper et al., 2002; Wang et al., 2017b; Xiao et al., 2014). Notably, a study of children 2 to 7 years showed higher GMV in the right STG in those with ASD, as compared to controls (Guo et al., 2021). The current findings add to this literature by showing this volumetric change in preterm newborns. On the other hand, while these findings suggest STG GMV as a risk marker (Mueller et al., 2013; Pierce, 2011), other brain regions, including those in the frontal and occipital lobes, have also been implicated in the pathophysiology of ASD (Courchesne et al., 2007; Guo et al., 2021; Liu et al., 2017; Sparks et al., 2002). The increase in regional GMV’s in preterm infants may reflect premature exposure to the external environment in utero and deficient synaptic pruning and/or compensatory overdevelopment of the brain. Notably, although characterized by early brain overgrowth (Bryńska, 2012; Courchesne et al., 2011; Ecker et al., 2015; Hazlett et al., 2017; Schumann et al., 2010), ASD may manifest stagnation in development later during puberty and early adulthood (Carper et al., 2002; Lange et al., 2015). Thus, age needs to be carefully considered in the characterization of structural brain abnormalities in ASD and likely other NDD.

### 4.2 Attention to facial features as reflected in eye-tracking metrics

Likewise, with the caveat, we discussed the current findings along with eye-tracking studies of ASD, because the bulk of this literature focused on ASD. Individuals with NDD exhibit attentional biases in social behaviors, as could be revealed in experimental studies of visual exploration of faces and social scenes with people engaged in conversation (Sadria et al., 2019; Shic et al., 2020). We observed more frequent of looking at the non-eyes in preterm, as compared to full-term toddlers, although the difference did not reach significance. To the extent that preterm birth represents a risk of ASD, this finding is consistent with previous reports of shorter and longer duration of gaze directed to eyes and mouths, respectively, in individuals with ASD viewing human faces. For instance, compared to the control group, two-year-olds with autism looked at others’ eyes significantly less (Jones and Klin, 2013) but at their mouths more (Jones et al., 2008b). At two years of age, children with ASD showed excessive mouths and reduced eyes gazing when observing social interactions that involved “dynamic” stimuli, such as child-oriented speech, or those that demanded shared attention (Campbell et al., 2014; Chawarska et al., 2012). In children around 5 years old shown 44 consecutive photographs of faces, those with ASD spent more time looking at mouths and less time looking at eyes, as compared to neurotypical children (Sadria et al., 2019). The current finding suggests that preferential looking at the mouths can be observed in toddlers, a very early stage of neurodevelopment, and that pre-term toddlers may acquire social information largely from mouth shape and movement (Jones et al., 2008a; Pelphrey et al., 2002; Wang et al., 2020).

Notably, earlier research pointed to the opposite conclusion in studies of adults (Corden et al., 2008; Falck-Ytter and von Hofsten, 2011; Klin et al., 2002). Thus, as with the imaging literature, age should be considered in understanding eye gazing pattern as a behavioral marker of ASD, and the current findings should be considered as specific to a sample 18 months of age. Possibly due to the accelerated brain development in the early years and delayed development later on, individuals with ASD have significantly higher and lower brain ages in early childhood and adolescence, respectively (He et al., 2020), and preterm infants share many of these features in their early years (Mendez et al., 2023), with similar facial recognition behaviors, such as looking more at mouths and less at eyes (Telford et al., 2016).

### 4.3 Potential sex differences

Previous studies documented higher frequency and severity of ASD in males than in females, as discussed earlier. Here, however, we observed indistinguishable Q-CHAT score between sexes, likely due to the small sample of PB. Earlier studies have also reported sex differences in attention and eye movement patterns during viewing of social scenes. For instance, females demonstrated more eye shifts amongst the facial areas, suggesting perhaps more effort in visual exploration, in neurotypical infants at 9 to 10 months (Rennels and Cummings, 2013). Chawarska and colleagues reported greater attention to social stimuli such as faces among girls at high risk for ASD (Chawarska et al., 2016). Throughout development, women pay more attention to faces than men (Gluckman and Johnson, 2013). Studies have also shown that girls relative to boys with ASD 6 to10 years looked at faces more (Harrop et al., 2018). Around age 1, males with ASD paid more attention to mouth emotions than male controls and females with ASD. In contrast, females with ASD had fewer gazes on mouth compared to female controls (Kleberg et al., 2019). Along with these earlier studies, we found sex differences in the current study; the earlier they were born, the more likely females but not males looked at non-eyes parts of the faces, which included largely the mouths. Kleberg et.al showed the opposite results in adults, with autistic men and women each looking more and less at the mouths relative to their neurotypical counterparts, again suggesting neurodevelopmental differences through the course of ASD (Kleberg et al., 2019).

### 4.4 Limitations and Conclusions

We considered a number of limitations of the study. First, the total Q-CHAT score did not show a significant difference between PB and RB or a correlation with any of the clinical or eye-tracking metrics, which we speculated may result from its low validity (<50%) (Thabtah and Peebles, 2019) or the relatively small sample size of PB. The latter also limits the possibility of a more thorough and powered investigation of sex differences. The subgroup sample of female PB in the database is small, and a larger dataset is needed to confirm these sex-different findings. Second, the dHCP removed speech from the eye-tracking task, which could have a significant influence on the behavioral outcomes. The current results should be considered as specific to this video stimulus. Third, mediation analysis suggests rSTG GMV as a mediator of greater frequency of face and mouth looking, seemingly antithetical in terms of the risk of social cognitive dysfunction. Thus, whether and to what extent elevated rSTG GMV reflects a risk phenotype or compensatory mechanism remains to be clarified (Hadad and Yashar, 2022). Fourth, the imaging data were collected within the first few weeks of life, and the eye-tracking data around eighteen months. Thus, many environmental factors, including parenting, can influence the findings beyond premature birth. A longitudinal study with imaging data collected at two or more time points would provide opportunities to charactering volumetric changes that may be crucial to social cognitive functioning during this early developmental stage. Finally, this research considered gray matter changes only; more work is needed to study white matter and functional changes (Dimond et al., 2019).

To conclude, preterm birth alters the gray matter volumes of right STG in newborns, with higher right STG GMV associated with eye-tracking marker of social cognitive function at 18 months, especially in females. Higher right STG GMV may represent a neural phenotype linking preterm birth to behavioral markers of social cognitive dysfunction in NDD, including ASD. Longitudinal studies will determine whether the volumetrics at birth and eye-tracking markers at 18 months would predict the onset and severity of ASD and other NDD.

## Data Availability

All data produced are available online at https://www.developingconnectome.org/

## Funding Support and Acknowledgement

The current study is supported by National Natural Science Foundation of China (12402350, U20A20388) and National Foreign Experts Program (H20240220). The developing Human Connectome Project was funded by the European Research Council under the European Union Seventh Framework Programme (FP/20072013)/ERC Grant Agreement no. 319456. This work was supported by the NIHR Biomedical Research Centres at Guys and St Thomas NHS Trust and the South London and Maudsley NHS Trust; the ESPRC/Wellcome Centre for Medical Engineering; and the MRC Centre for Neurodevelopmental Disorders. TA was supported by MRC Clinician Scientist Fellowship (MR/P008712/1) and MRC translation support award (MR/V036874/1). JO’M was supported by a Sir Henry Dale Fellowship jointly funded by the Wellcome Trust and the Royal Society (Grant Number 206675/Z/17/Z).

## Author contributions

I. Conception and design: Guangfei Li and Chiang-Shan R. Li
II. Administrative support: Dongmei Hao and Chiang-Shan R. Li
III. Provision of study materials or patients: Yanan Su, Yiyao Ye-Lin, Xiaolin Wang, Ruolin Zhang, Lin Yang and Chiang-Shan R. Li
IV. Collection and assembly of data: Yanan Su, Shanmei Wang and Guangfei Li
V. Data analysis and interpretation: Yanan Su, Guangfei Li, Clara S. Li and Chiang-Shan R. Li.
VI. Manuscript writing: Yanan Su, Guangfei Li and Chiang-Shan R. Li.
VII. Final approval of manuscript: All authors.

## Ethics Statement

The authors are accountable for all aspects of the work in ensuring that questions related to the accuracy or integrity of any part of the work are appropriately investigated and resolved. The study was conducted in accordance with the Declaration of Helsinki (as revised in 2013) and approved by United Kingdom Health Research Authority (Research Ethics Committee reference number: 14/LO/1169). Written informed consent to participate in this study was provided by the participants’ legal guardian/next of kin.

## Data Availability Statemen**t**

The developing HCP data are available to the public at http://www.developingconnectome.org/data-release/third-data-release/, or https://nda.nih.gov/edit_collection.html?id=3955

## Supplement

### A. Behavioral markers of ASD

Children with ASD are usually identified around 3 to 4 years of age, despite earlier signs of atypical development (Lai et al., 2014). Assessment of very young children for risks of ASD is challenging. Many children later diagnosed with ASD do not differ from typically developing children in social smiling and facial gaze at 6 and 12 months of age (Landa and Garrett-Mayer, 2006). In addition to clinical assessments, such as Denver Developmental Screening Test (De-Andrés-Beltrán et al., 2015), Bayley Infant Neurodevelopmental Screener (Albers and Grieve, 2007), Quantitative Checklist for Autism in Toddlers (Allison et al., 2008), and Modified Checklist for Autism in Toddlers (Dereu, 2021), tracking of eye movements during viewing of social scenes can help in identifying behavioral markers of ASD in very young children (Murias et al., 2018; Shan et al., 2023).

### B. Supplementary Tables and Results

**Supplementary Table S1.**
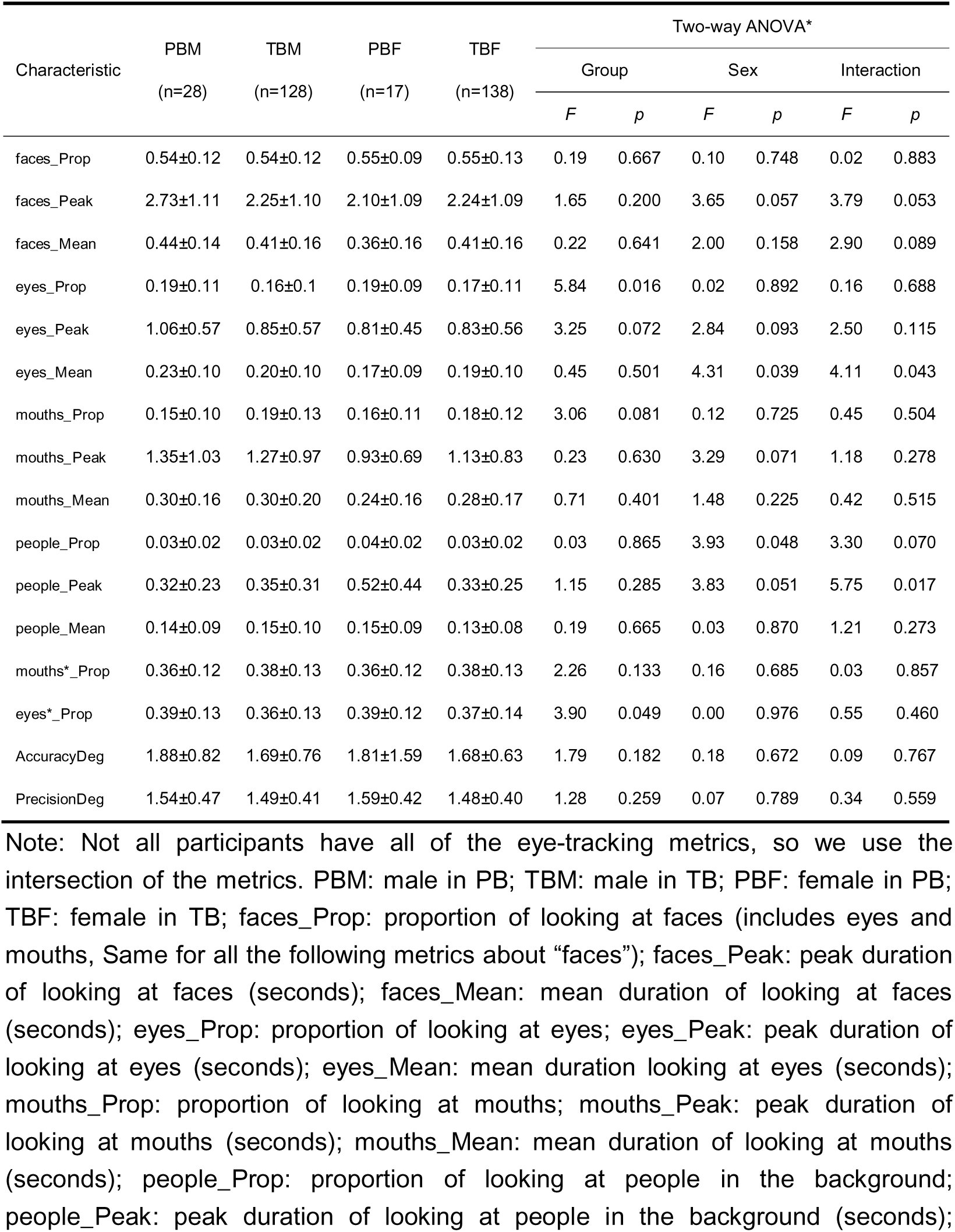
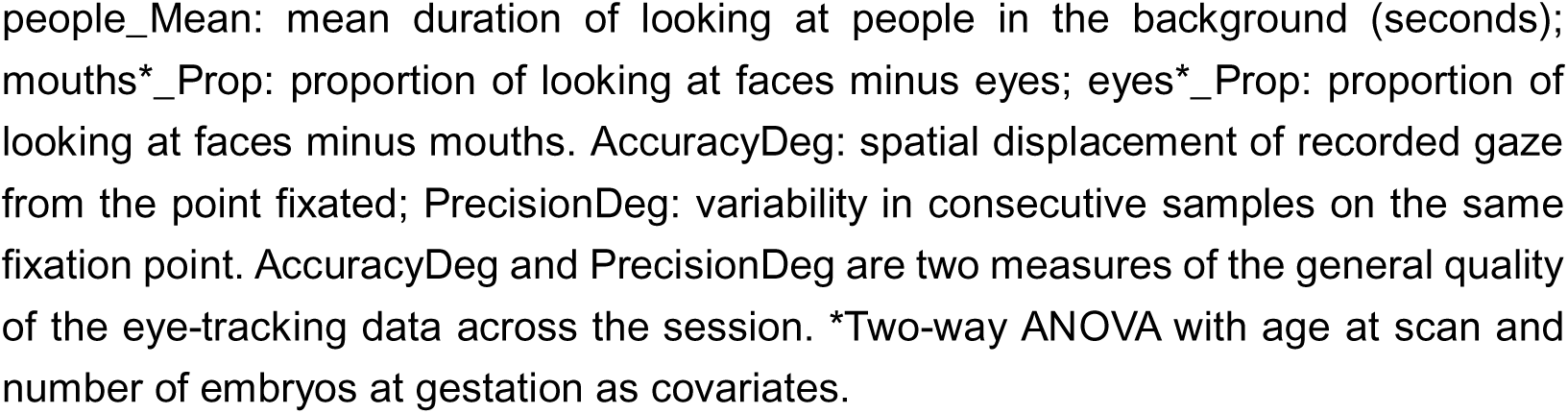
All Eye-tracking measures of the participants.

#### PB vs. TB in clinical and eye-tracking metrics

Clinical characteristics as well as the statistics of group x sex ANOVA are summarized in **Table 1**, age at birth showed significant group (F = 1417, p < 0.001) and sex (F = 10.17, p = 0.002) main as well as interaction (F = 12.19, p = 0.001) effect. In planned comparisons, PB showed a younger age at birth than TB (PB: 32.24 ± 3.36; TB: 40.05 ± 1.26), as expected. Female were born later than male (Female: 39.07 ± 3.23; Male: 38.95 ± 3.09). PBF showed an older age at birth than PBM (PBF: 31.38 ± 3.23; PBM: 32.80 ± 3.36) and showed a younger age at birth than TBF (PBF: 31.38 ± 3.23; TBF: 40.08 ± 1.30). TBM were born later than PBM (PBM: 32.80 ± 3.36; TBM: 40.02 ± 1.24) and were born earlier than TBF (TBF: 40.27 ± 1.23; TBM: 40.05 ± 1.26).

Age at scan showed a significant sex (F = 4.60, p = 0.032) main effect with females scanned later than males (Female: 41.56 ± 1.80; Male: 41.19 ± 1.72) but not group main (F = 2.90, p = 0.089) or interaction (F = 0.53, p = 0.465) effect. Age at eye-tracking experiment showed a significant group (F = 29.96, p < 0.001) but not sex (F = 0.52, p =0.472) main or interaction (F = 0.38, p = 0.538) effect. PB performed eye-tracking experiments at a later age than TB (PB: 21.27 ± 2.70; TB: 19.01 ± 1.97).

Mo_edu_age showed no group (F < 0.01, p =0.986) or sex (F = 1.00, p = 0.317) main or interaction (F = 0.86, p =0.355) effect. Fa_edu_age showed significant sex (F = 6.84, p = 0.009) but not group (F = 3.34, p = 0.068) main or interaction effect (F = 2.34, p = 0.127). Fathers of PB (PB: 21.84 ± 3.14; TB: 23.46 ± 4.86) had a younger age when they were last in continuous full-time education.

TIV and “singleton” had significant group main (TIV: F = 49.26, p < 0.001; Singleton: F = 94.34, p < 0.001), but not sex main (TIV: F = 0.33, p = 0.564; Singleton: F = 0.23, p = 0.634) or interaction (TIV: F = 0.35, p = 0.553; Singleton: F = 0.43, p = 0.512) effect. TB had a greater TIV than PB (PB: 385.66 ± 8.84; TB: 392.12 ± 7.02), and PB were more likely to be born multiple births (PB: 27/76; TB: 20/493). Female showed a smaller TIV than male (Female: 390.87 ± 7.66; Male: 391.59 ± 7.55), and male were more likely to be born multiple births (Female: 20/259; Male: 27/310).

Radiology score (RS) showed significant group main effect (F = 5.79, p = 0.016) and interaction effect (F = 3.93, p = 0.048), but not sex main effect (F = 0.04, p = 0.842). More children in PB had a RS of 2 than those in TB (PB: 37/76; TB: 177/493). PBF showed a worse RS than PBM (PBF: 17/30; PBM: 20/46) and showed also a worse RS than TBF (PBF: 17/30; TBF: 69/229). TBM had a better RS than PBM (PBM: 20/46; TBM: 108/264) and had a worse RS than TBF (TBF: 69/229; TBM: 108/264).

**Supplementary Table S2.** Correlation of Q-CHAT score and age at birth with eye-tracking measures.

| Characteristic |  | All<br>(n = 311) | Male<br>(n = 156) | Female<br>(n = 155) | PB<br>(n = 45) | TB<br>(n = 266) | PBM<br>(n = 28) | TBM<br>(n = 128) | PBF<br>(n = 17) | TBF<br>(n = 138) |
| --- | --- | --- | --- | --- | --- | --- | --- | --- | --- | --- |
| *Q-CHAT vs. birth age and eye-tracking measures* |  |  |  |  |  |  |  |  |  |  |
| birth_age | *r* | 0.03 | 0.14 | -0.08 | -0.14 | 0.06 | -0.02 | 0.18 | -0.22 | -0.05 |
|  | *p* | 0.602 | 0.096 | 0.323 | 0.397 | 0.314 | 0.916 | 0.044* | 0.447 | 0.597 |
| faces_Prop | *r* | -0.03 | 0.04 | -0.10 | -0.21 | 0.00 | -0.29 | 0.13 | -0.09 | -0.12 |
|  | *p* | 0.659 | 0.601 | 0.217 | 0.186 | 0.965 | 0.155 | 0.155 | 0.754 | 0.162 |
| mouths*_Prop | *r* | 0.02 | 0.10 | -0.08 | -0.05 | 0.02 | -0.12 | 0.14 | -0.08 | -0.10 |
|  | *p* | 0.796 | 0.222 | 0.325 | 0.752 | 0.732 | 0.567 | 0.128 | 0.775 | 0.235 |
| eyes*_Prop | *r* | -0.06 | -0.07 | -0.04 | -0.23 | -0.16 | -0.31 | 0.02 | 0.04 | -0.04 |
|  | *p* | 0.340 | 0.391 | 0.643 | 0.154 | 0.800 | 0.131 | 0.857 | 0.893 | 0.633 |

| *birth_age vs. eye-tracking measures* |  |  |  |  |  |  |  |  |  |  |
| --- | --- | --- | --- | --- | --- | --- | --- | --- | --- | --- |
| faces_Prop | *r* | -0.02 | 0.03 | -0.05 | -0.03 | 0.03 | 0.12 | 0.16 | -0.29 | -0.08 |
|  | *p* | 0.753 | 0.685 | 0.517 | 0.841 | 0.665 | 0.570 | 0.071 | 0.307 | 0.382 |
| mouths*_Prop | *r* | 0.03 | 0.14 | -0.04 | -0.23 | 0.01 | 0.12 | -0.10 | -0.67 | -0.08 |
|  | *p* | 0.557 | 0.094 | 0.631 | 0.140 | 0.928 | 0.567 | 0.279 | 0.009* | 0.372 |
| eyes*_Prop | *r* | -0.05 | -0.11 | -0.01 | 0.25 | -0.01 | 0.06 | 0.11 | 0.48 | -0.05 |
|  | *p* | 0.348 | 0.165 | 0.915 | 0.122 | 0.836 | 0.767 | 0.237 | 0.081 | 0.538 |

Note: Not all participants have all of the eye-tracking metrics, so we use the intersection of the metrics. PBM: male in PB; TBM: male in TB; PBF: female in PB; TBF: female in TB; birth\_age: gestational age at birth; faces\_Prop: proportion of looking at faces; mouths\*\_Prop: proportion of looking at faces minus eyes; eyes\*\_Prop: proportion of looking at faces minus mouths. With age at scan, TIV, sex and number of embryos at gestation as covariates in All, PB and TB; with age at scan and number of embryos at gestation as covariates in other groups. \*p<0.05, uncorrected.

**Supplementary Table S3.**
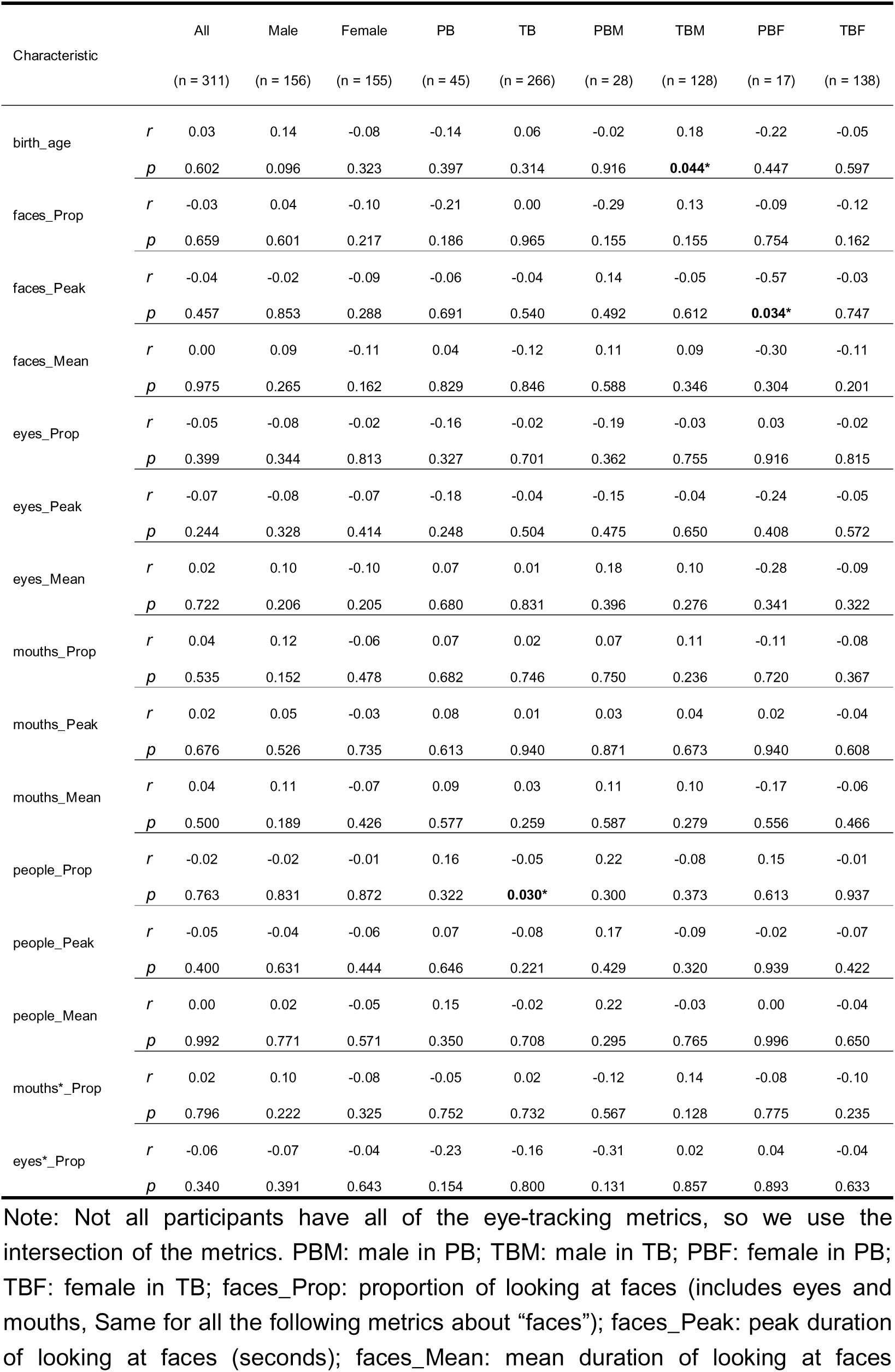
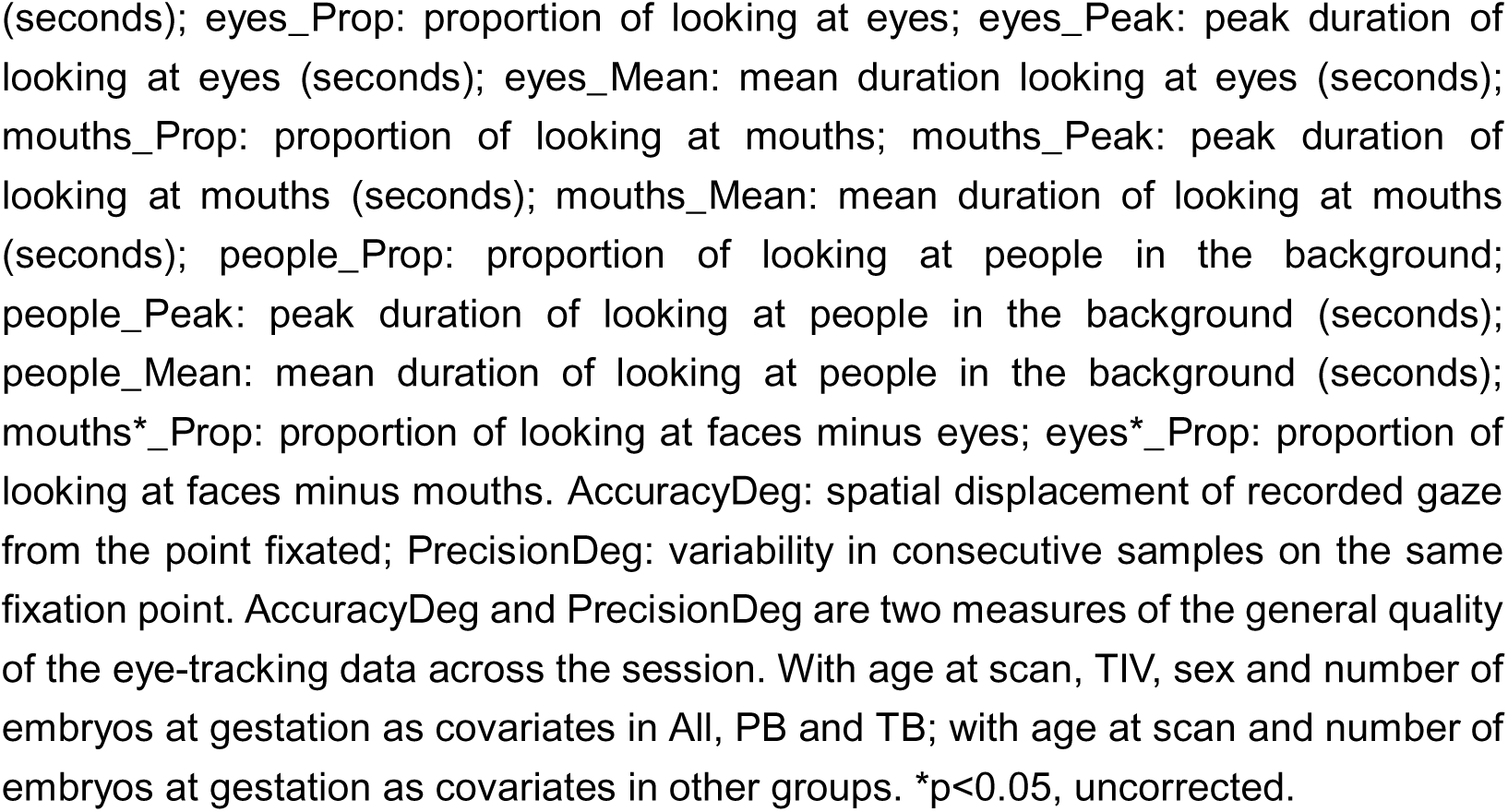
Correlation of Q-CHAT score with age at birth and eye-tracking measures.

**Supplementary Table S4.**
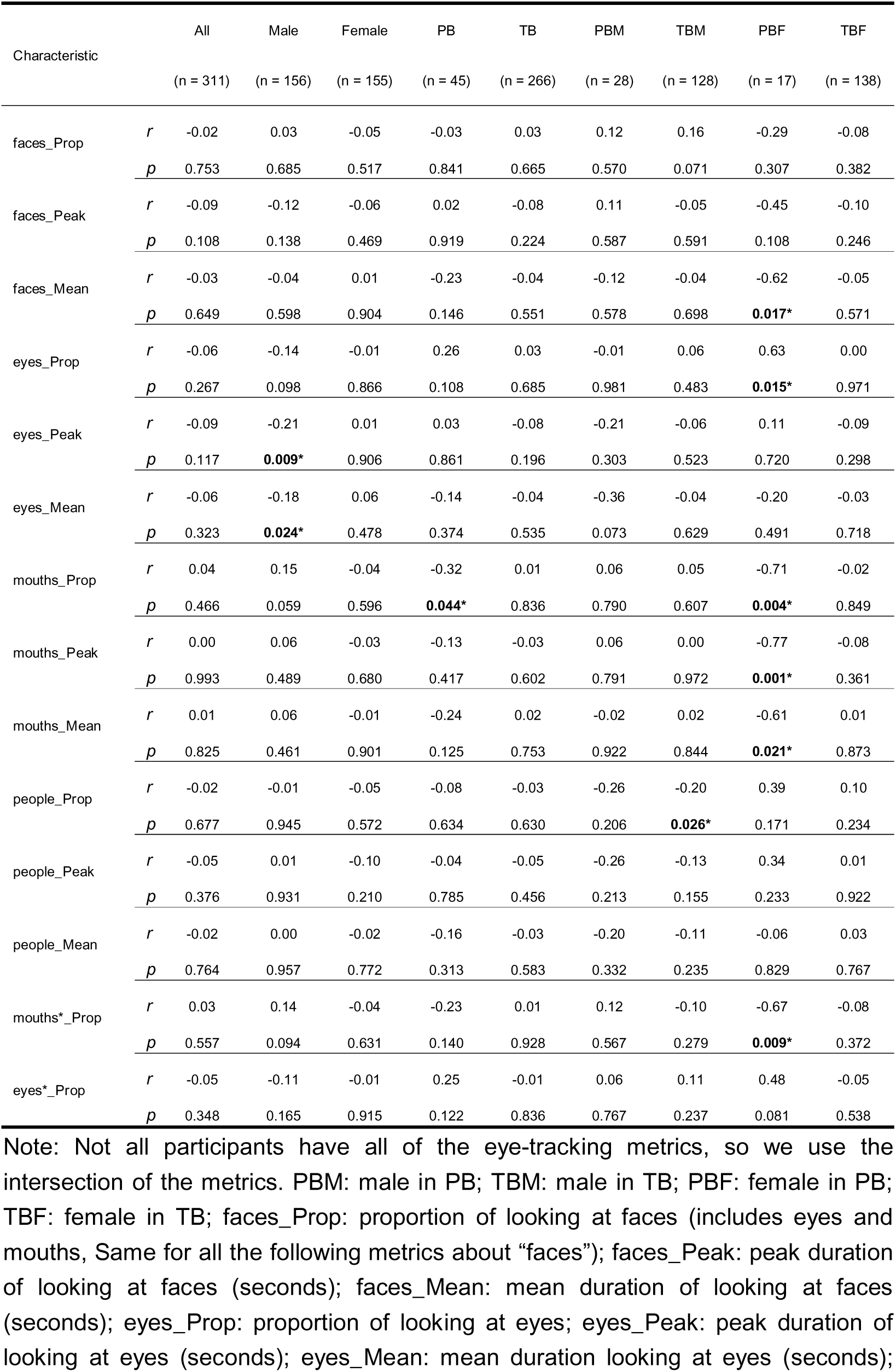
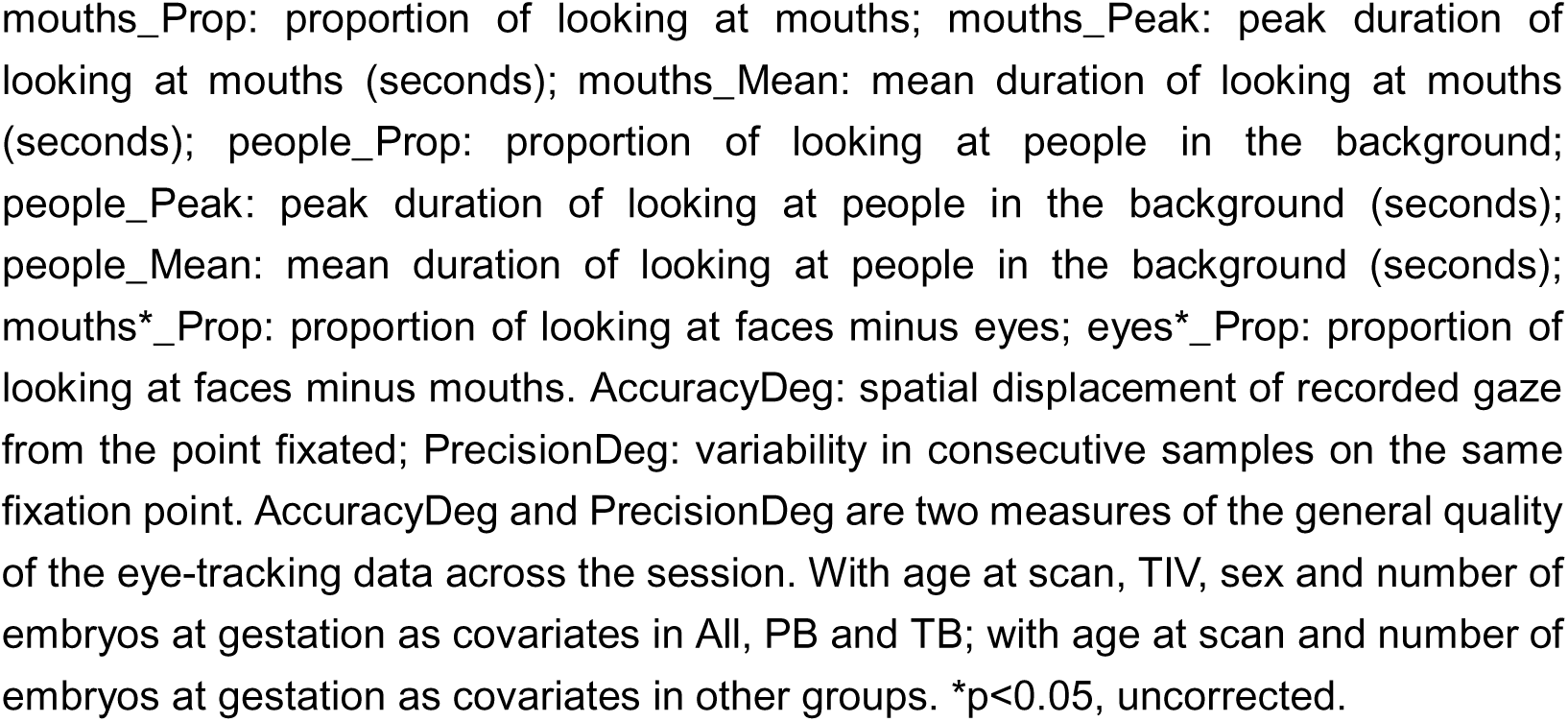
Correlation of age at birth and eye-tracking measures.

**Supplementary Table S5.** Correlation of right and left STG GMV with birth_age and three eye-tracking metrics.

|  |  | All | Male | Female | PB | TB | PBM | TBM | PBF | TBF |
| --- | --- | --- | --- | --- | --- | --- | --- | --- | --- | --- |
| Characteristic |  | (n = 311) | (n = 156) | (n = 155) | (n = 45) | (n = 266) | (n = 28) | (n = 128) | (n = 17) | (n = 138) |
| *Right STG GMV and eye-tracking measures* |  |  |  |  |  |  |  |  |  |  |
| birth_age | *r* | -0.47 | -0.42 | -0.51 | -0.37 | -0.03 | -0.23 | -0.01 | -0.55 | -0.03 |
|  | *p* | **0.000** | **0.000** | **0.000** | 0.017 | 0.660 | 0.276 | 0.941 | 0.040 | 0.749 |
| faces_Prop | *r* | 0.10 | 0.12 | 0.08 | 0.50 | 0.08 | 0.51 | 0.05 | 0.42 | 0.10 |
|  | *p* | 0.070 | 0.153 | 0.319 | **0.001** | 0.273 | 0.009 | 0.568 | 0.131 | 0.245 |
| mouths*_Prop | *r* | 0.01 | 0.02 | -0.00 | 0.39 | 0.03 | 0.32 | 0.04 | 0.457 | 0.03 |
|  | *p* | 0.821 | 0.811 | 0.983 | 0.012 | 0.631 | 0.125 | 0.654 | 0.101 | 0.718 |
| eyes*_Prop | *r* | 0.12 | 0.15 | 0.09 | 0.18 | 0.07 | 0.34 | 0.03 | -0.14 | 0.09 |
|  | *p* | 0.037 | 0.071 | 0.269 | 0.273 | 0.266 | 0.098 | 0.747 | 0.643 | 0.277 |
| *Left STG GMV vs. eye-tracking measures* |  |  |  |  |  |  |  |  |  |  |
| birth_age | *r* | -0.33 | -0.19 | -0.44 | -0.16 | -0.07 | 0.05 | 0.01 | -0.48 | -0.12 |
|  | *p* | **0.000** | 0.021 | **0.000** | 0.306 | 0.257 | 0.815 | 0.898 | 0.082 | 0.164 |
| faces_Prop | *r* | 0.18 | 0.13 | 0.22 | 0.41 | 0.15 | 0.45 | 0.04 | 0.40 | 0.24 |
|  | *p* | 0.002 | 0.120 | 0.007 | 0.007 | 0.014 | 0.025 | 0.646 | 0.155 | 0.005 |
| mouths*_Prop | *r* | 0.04 | 0.03 | 0.04 | 0.19 | 0.04 | 0.24 | 0.02 | 0.07 | 0.07 |
|  | *p* | 0.529 | 0.679 | 0.652 | 0.225 | 0.474 | 0.251 | 0.836 | 0.821 | 0.429 |
| eyes*_Prop | *r* | 0.22 | 0.142 | 0.29 | 0.27 | 0.20 | 0.29 | 0.06 | 0.34 | 0.31 |
|  | *p* | **0.000** | 0.080 | **0.000** | 0.094 | **0.001** | 0.156 | 0.543 | 0.239 | **0.000** |

With age at scan, TIV, sex and number of embryos at gestation as covariates in All, PB and TB; with age at scan and number of embryos at gestation as covariates in other groups. We employed a p value of $0.05/(2 \text{ regions} \times 3 \text{ metrics}) = 0.008$ as the threshold.

**Supplementary Table S6a.**
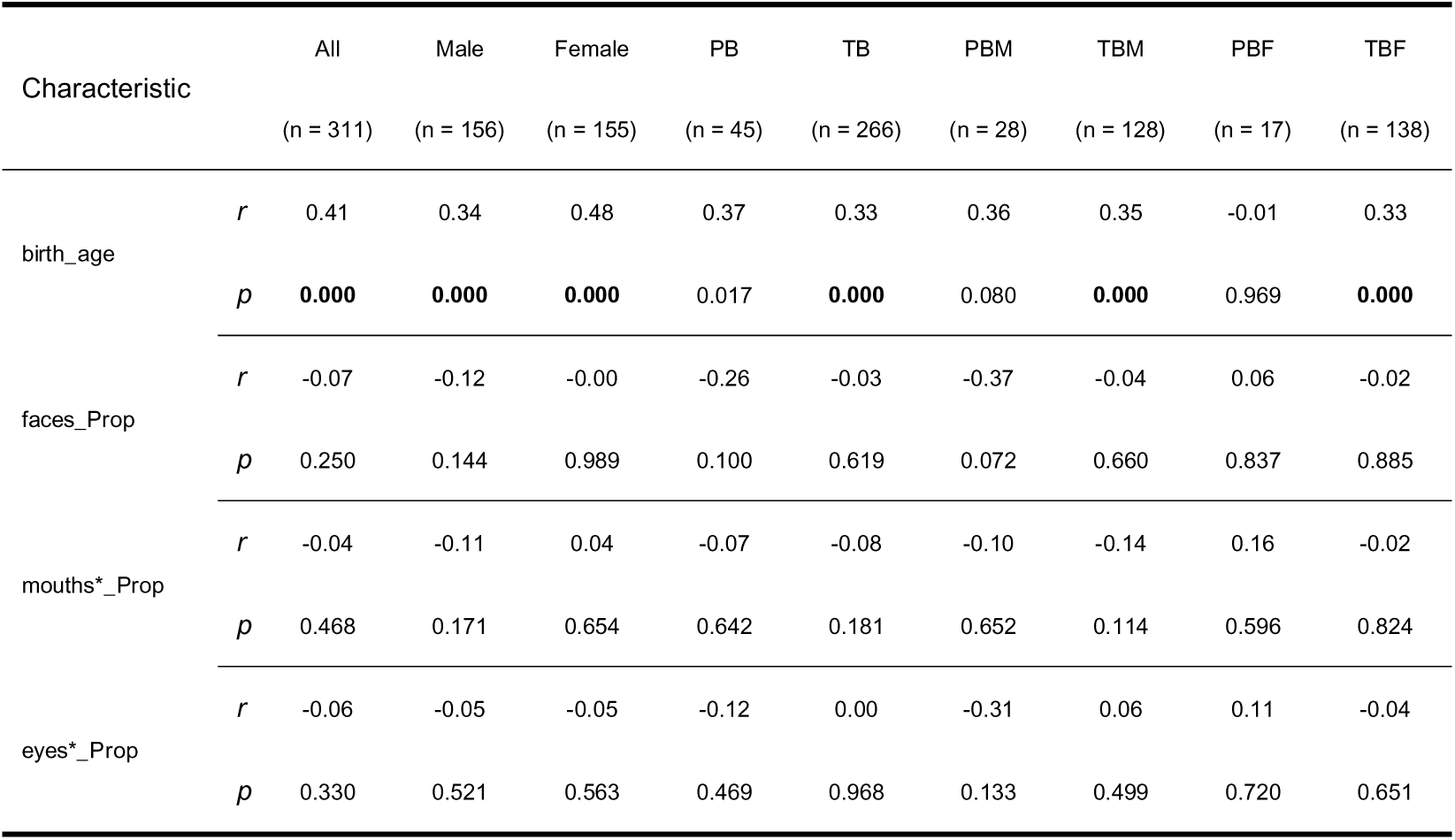
Correlation of **HIP_L** with birth_age and three eye-tracking metrics.

| Characteristic |  | All<br>(n = 311) | Male<br>(n = 156) | Female<br>(n = 155) | PB<br>(n = 45) | TB<br>(n = 266) | PBM<br>(n = 28) | TBM<br>(n = 128) | PBF<br>(n = 17) | TBF<br>(n = 138) |
| --- | --- | --- | --- | --- | --- | --- | --- | --- | --- | --- |
| birth_age | *r* | 0.41 | 0.34 | 0.48 | 0.37 | 0.33 | 0.36 | 0.35 | -0.01 | 0.33 |
|  | *p* | **0.000** | **0.000** | **0.000** | 0.017 | **0.000** | 0.080 | **0.000** | 0.969 | **0.000** |
| faces_Prop | *r* | -0.07 | -0.12 | -0.00 | -0.26 | -0.03 | -0.37 | -0.04 | 0.06 | -0.02 |
|  | *p* | 0.250 | 0.144 | 0.989 | 0.100 | 0.619 | 0.072 | 0.660 | 0.837 | 0.885 |
| mouths*_Prop | *r* | -0.04 | -0.11 | 0.04 | -0.07 | -0.08 | -0.10 | -0.14 | 0.16 | -0.02 |
|  | *p* | 0.468 | 0.171 | 0.654 | 0.642 | 0.181 | 0.652 | 0.114 | 0.596 | 0.824 |
| eyes*_Prop | *r* | -0.06 | -0.05 | -0.05 | -0.12 | 0.00 | -0.31 | 0.06 | 0.11 | -0.04 |
|  | *p* | 0.330 | 0.521 | 0.563 | 0.469 | 0.968 | 0.133 | 0.499 | 0.720 | 0.651 |

**Supplementary Table S6b.** Correlation of **HIP_R** with birth_age and three eye-tracking metrics.

| Characteristic |  | All<br>(n = 311) | Male<br>(n = 156) | Female<br>(n = 155) | PB<br>(n = 45) | TB<br>(n = 266) | PBM<br>(n = 28) | TBM<br>(n = 128) | PBF<br>(n = 17) | TBF<br>(n = 138) |
| --- | --- | --- | --- | --- | --- | --- | --- | --- | --- | --- |
| birth_age | *r* | 0.42 | 0.33 | 0.49 | 0.29 | 0.31 | 0.19 | 0.35 | 0.05 | 0.32 |
|  | *p* | **0.000** | **0.000** | **0.000** | 0.068 | **0.000** | 0.357 | **0.000** | 0.878 | **0.000** |
| faces_Prop | *r* | -0.03 | -0.09 | 0.06 | -0.10 | 0.00 | -0.23 | -0.02 | 0.37 | 0.04 |
|  | *p* | 0.660 | 0.246 | 0.440 | 0.514 | 0.945 | 0.262 | 0.801 | 0.198 | 0.632 |
| mouths*_Prop | *r* | 0.01 | -0.05 | 0.10 | -0.08 | -0.02 | -0.08 | -0.08 | 0.15 | 0.07 |
|  | *p* | 0.840 | 0.517 | 0.210 | 0.632 | 0.771 | 0.708 | 0.386 | 0.608 | 0.431 |
| eyes*_Prop | *r* | -0.04 | -0.07 | 0.00 | 0.08 | 0.00 | -0.20 | 0.03 | 0.45 | -0.03 |
|  | *p* | 0.490 | 0.375 | 0.997 | 0.611 | 0.963 | 0.333 | 0.731 | 0.105 | 0.775 |

**Supplementary Table S6c.** Correlation of **REC_L** with birth_age and three eye-tracking metrics.

| Characteristic |  | All<br>(n = 311) | Male<br>(n = 156) | Female<br>(n = 155) | PB<br>(n = 45) | TB<br>(n = 266) | PBM<br>(n = 28) | TBM<br>(n = 128) | PBF<br>(n = 17) | TBF<br>(n = 138) |
| --- | --- | --- | --- | --- | --- | --- | --- | --- | --- | --- |
| birth_age | *r* | 0.43 | 0.37 | 0.50 | 0.43 | 0.10 | 0.24 | 0.18 | 0.58 | 0.04 |
|  | *p* | **0.000** | **0.000** | **0.000** | 0.004 | 0.122 | 0.245 | 0.051 | 0.029 | 0.632 |
| faces_Prop | *r* | -0.08 | -0.10 | -0.04 | -0.42 | -0.02 | -0.44 | 0.01 | -0.20 | -0.04 |
|  | *p* | 0.183 | 0.226 | 0.590 | 0.006 | 0.712 | 0.030 | 0.889 | 0.491 | 0.672 |
| mouths*_Prop | *r* | -0.03 | 0.02 | -0.08 | -0.27 | -0.05 | -0.06 | 0.01 | -0.42 | -0.07 |
|  | *p* | 0.575 | 0.791 | 0.338 | 0.08 | 0.467 | 0.771 | 0.930 | 0.137 | 0.417 |
| eyes*_Prop | *r* | -0.06 | -0.16 | 0.05 | -0.16 | 0.02 | -0.45 | -0.01 | 0.30 | 0.05 |
|  | *p* | 0.322 | 0.055 | 0.527 | 0.309 | 0.811 | 0.023 | 0.890 | 0.297 | 0.595 |

**Supplementary Table S6d.** . Correlation of **ROL_L** with birth_age and three eye-tracking metrics.

| Characteristic |  | All<br>(n = 311) | Male<br>(n = 156) | Female<br>(n = 155) | PB<br>(n = 45) | TB<br>(n = 266) | PBM<br>(n = 28) | TBM<br>(n = 128) | PBF<br>(n = 17) | TBF<br>(n = 138) |
| --- | --- | --- | --- | --- | --- | --- | --- | --- | --- | --- |
| birth_age | *r* | 0.42 | 0.28 | 0.52 | 0.23 | 0.36 | -0.01 | 0.28 | 0.12 | 0.42 |
|  | *p* | **0.000** | **0.000** | **0.000** | 0.150 | **0.000** | 0.977 | 0.002 | 0.691 | **0.000** |
| faces_Prop | *r* | -0.06 | -0.06 | -0.05 | -0.40 | 0.02 | -0.39 | 0.10 | -0.29 | -0.03 |
|  | *p* | 0.268 | 0.454 | 0.533 | 0.008 | 0.750 | 0.057 | 0.254 | 0.320 | 0.770 |
| mouths*_Prop | *r* | 0.02 | -0.00 | 0.05 | -0.20 | 0.03 | -0.19 | 0.03 | 0.04 | 0.05 |
|  | *p* | 0.753 | 0.967 | 0.558 | 0.199 | 0.605 | 0.355 | 0.756 | 0.886 | 0.594 |
| eyes*_Prop | *r* | -0.14 | -0.09 | -0.16 | -0.29 | -0.06 | -0.37 | 0.09 | -0.31 | -0.15 |
|  | *p* | 0.018 | 0.249 | 0.056 | 0.062 | 0.319 | 0.069 | 0.341 | 0.274 | 0.077 |

**Supplementary Table S6e.** Correlation of **ROL_R** with birth_age and three eye-tracking metrics.

| Characteristic |  | All<br>(n = 311) | Male<br>(n = 156) | Female<br>(n = 155) | PB<br>(n = 45) | TB<br>(n = 266) | PBM<br>(n = 28) | TBM<br>(n = 128) | PBF<br>(n = 17) | TBF<br>(n = 138) |
| --- | --- | --- | --- | --- | --- | --- | --- | --- | --- | --- |
| birth_age | *r* | 0.47 | 0.39 | 0.56 | 0.40 | 0.38 | 0.24 | 0.38 | 0.46 | 0.40 |
|  | *p* | **0.000** | **0.000** | **0.000** | 0.008 | **0.000** | 0.258 | **0.000** | 0.102 | **0.000** |
| faces_Prop | *r* | -0.07 | -0.09 | -0.02 | -0.18 | -0.03 | -0.17 | -0.00 | 0.03 | -0.04 |
|  | *p* | 0.237 | 0.259 | 0.822 | 0.242 | 0.564 | 0.412 | 0.969 | 0.912 | 0.658 |
| mouths*_Prop | *r* | 0.01 | -0.01 | 0.07 | -0.20 | 0.01 | -0.13 | -0.01 | -0.18 | 0.06 |
|  | *p* | 0.818 | 0.900 | 0.422 | 0.217 | 0.825 | 0.550 | 0.940 | 0.539 | 0.475 |
| eyes*_Prop | *r* | -0.11 | -0.13 | -0.09 | 0.01 | -0.09 | -0.16 | -0.02 | 0.41 | -0.15 |
|  | *p* | 0.047 | 0.114 | 0.266 | 0.976 | 0.135 | 0.439 | 0.800 | 0.151 | 0.087 |

**Supplementary Table S7.**
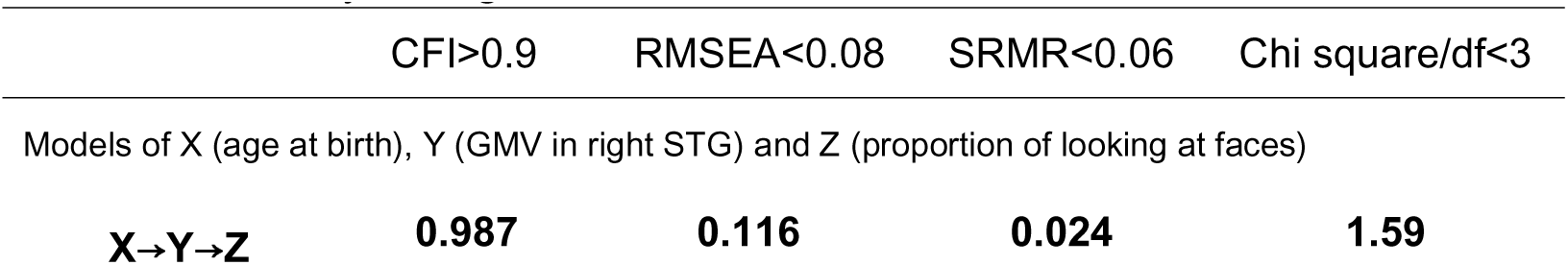
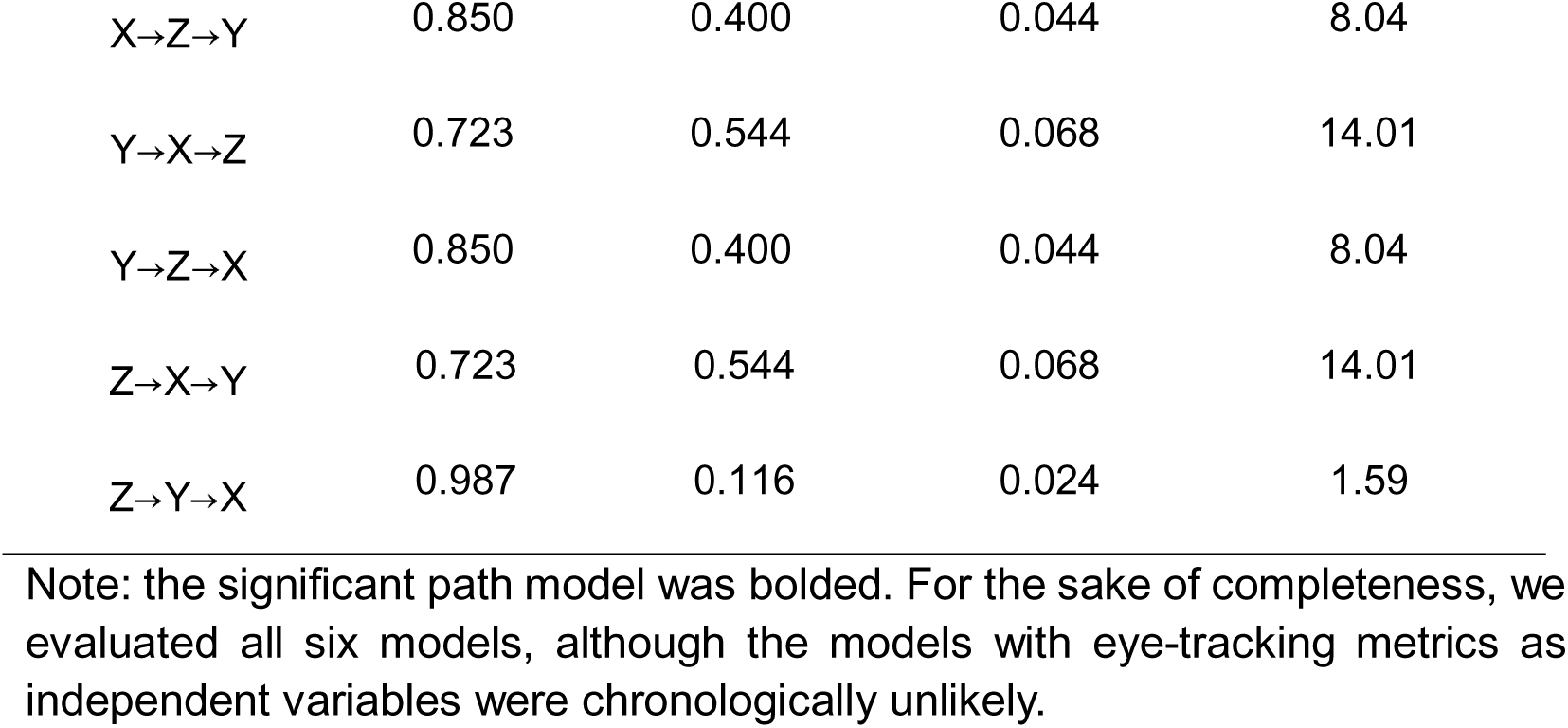
Statistics of path analyses of age at birth, GMV in right STG and proportion of looking at faces, with age at scan, sex, TIV, and number of embryos at gestation as covariates.

**Supplementary Tables S8.** Statistics of path analyses of age at birth, GMV in right STG and proportion of looking at mouths*, with age at scan, sex, TIV, and number of embryos at gestation as covariates.

|  | CFI>0.9 | RMSEA<0.08 | SRMR<0.06 | Chi square/df<3 |
| --- | --- | --- | --- | --- |
| Models of X (age at birth), Y (GMV in right STG) and Z (proportion of looking at mouths*) |  |  |  |  |
| **X→Y→Z** | **1.000** | **0.000** | **0.014** | **0.44** |
| X→Z→Y | 0.908 | 0.284 | 0.035 | 4.55 |
| Y→X→Z | 0.888 | 0.314 | 0.044 | 5.33 |
| Y→Z→X | 0.908 | 0.284 | 0.035 | 4.55 |
| Z→X→Y | 0.888 | 0.314 | 0.044 | 5.33 |
| Z→Y→X | 1.000 | 0.000 | 0.014 | 0.44 |

Note: the significant path model was bolded. For the sake of completeness, we evaluated all six models, although the models with eye-tracking metrics as independent variables were chronologically unlikely.

### C. Supplementary Figures

**Supplementary Figure S1.**
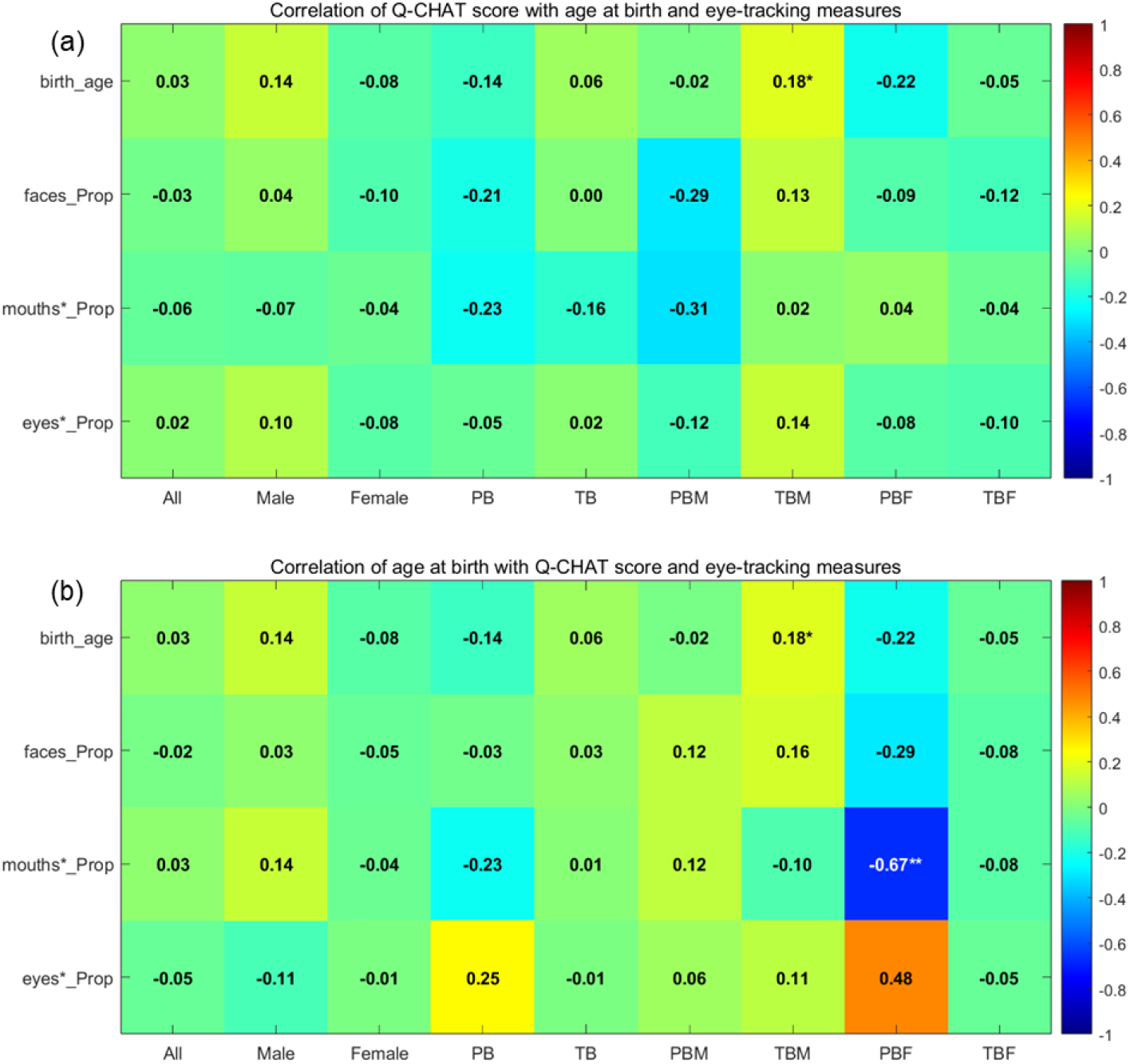
Correlation matrices based on **Table 3**. PBM: male in PB; TBM: male in TB; PBF: female in PB; TBF: female in TB; birth_age: gestational age at birth; faces_Prop: proportion of looking at faces; mouths*_Prop: proportion of looking at faces minus eyes; eyes*_Prop: proportion of looking at faces minus mouths. With age at scan, sex and number of embryos at gestation as covariates in All, PB and TB; with age at scan and number of embryos at gestation as covariates in other groups. *p<0.05; **p<0.01.

**Supplementary Figure S2.**
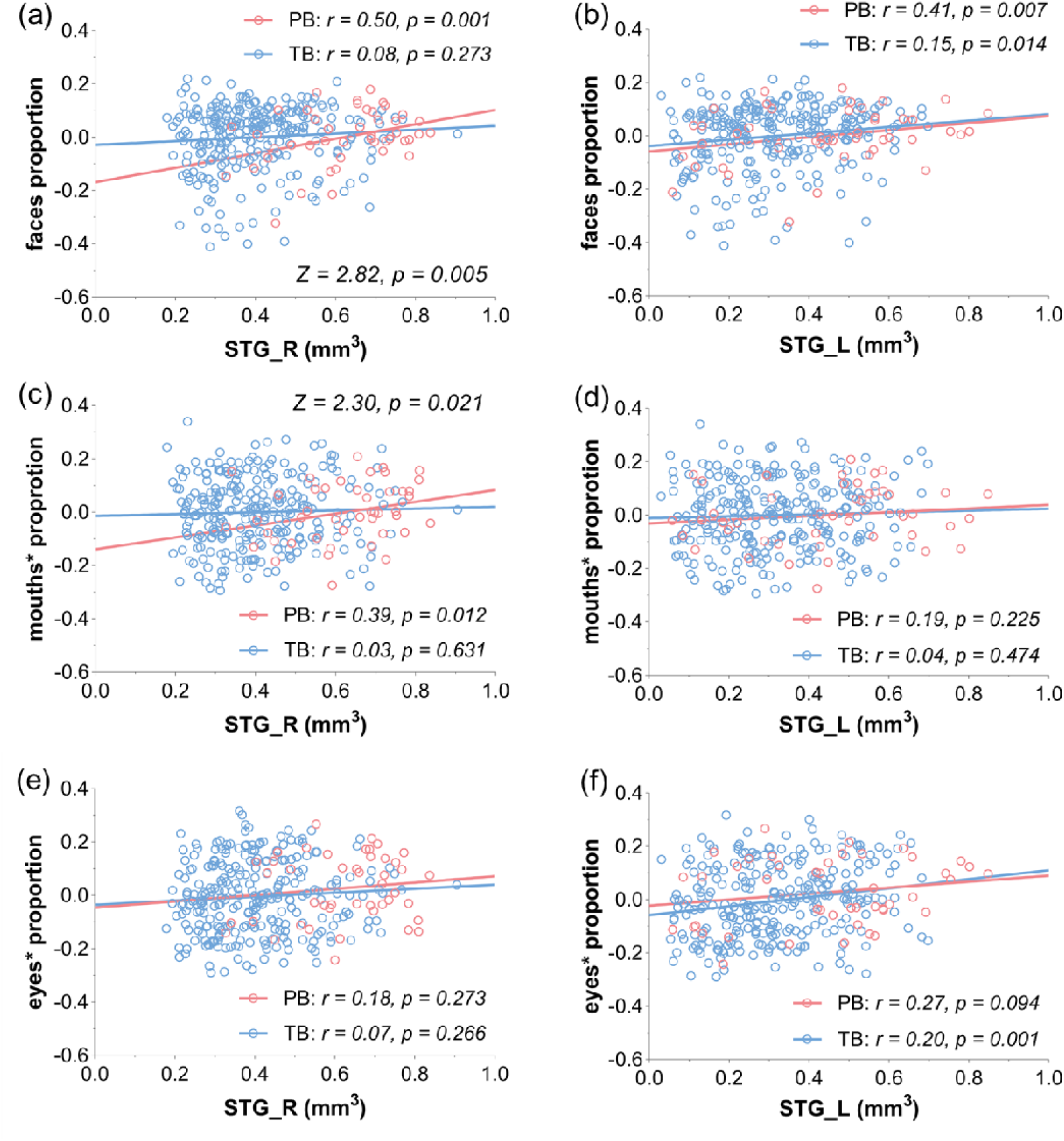
Regression of right STG (left column) and left STG (right column) with the three primary eye-tracking metrics. PB in red and TB in blue. The Z and p values of slope tests are shown for those regressions that differed significantly in slope between PB and TB.

**Supplementary Figure S3.**
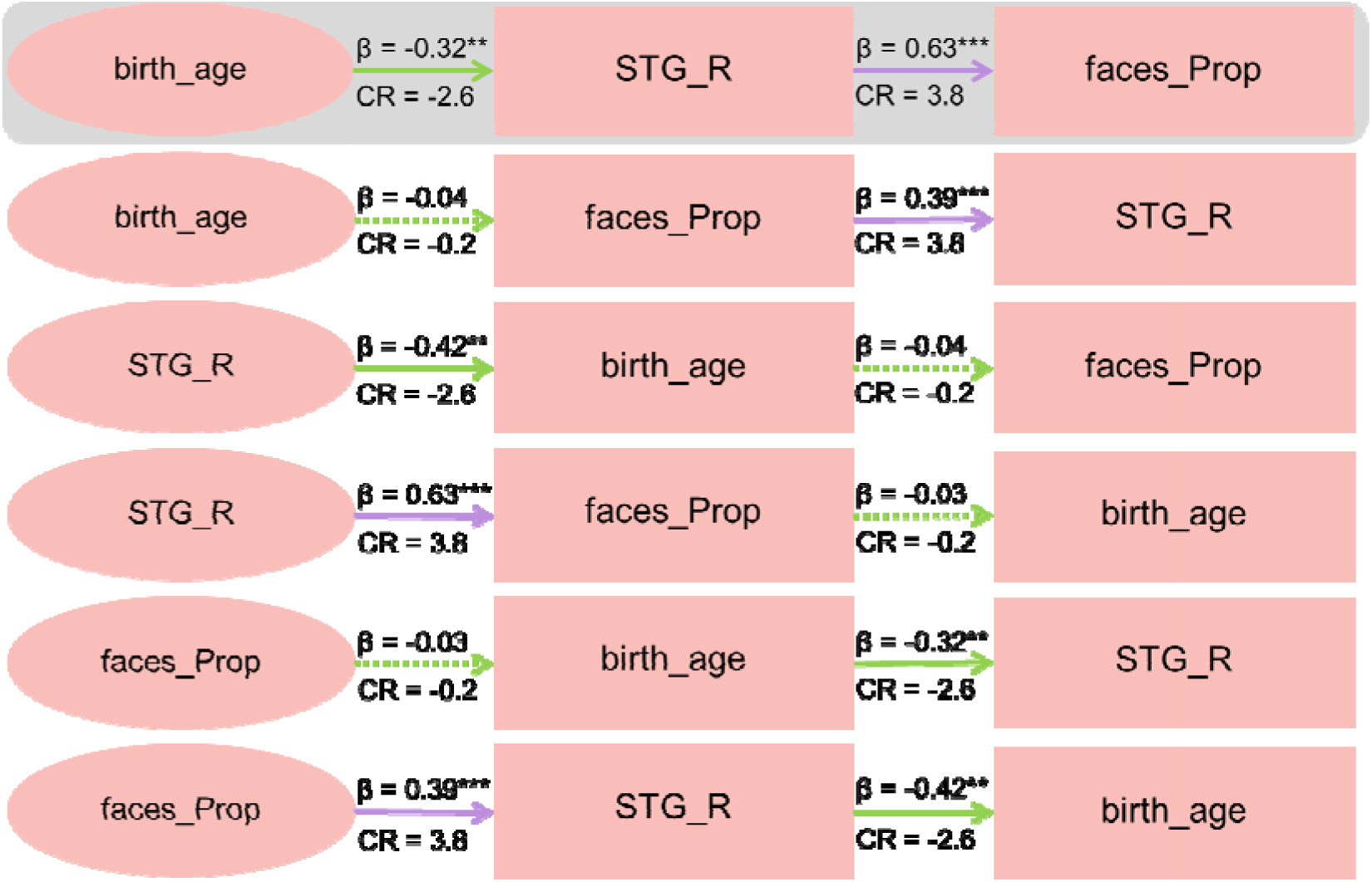
Path analysis in PB of age at birth, right STG GMV, and proportion of looking at faces. Solid line and dotted line each indicates a significant and non-significant path effect. Purple and green arrow each indicates positive and negative effect. The modeling included age at scan, TIV, sex, and the number of embryos at gestation as covariates. **p<0.01, ***p<0.001. CR: critical ratio. Note: For the sake of completeness, we evaluated all six models, although the models with proportion of looking at faces as independent variables were chronologically unlikely.

**Supplementary Figure S4.**
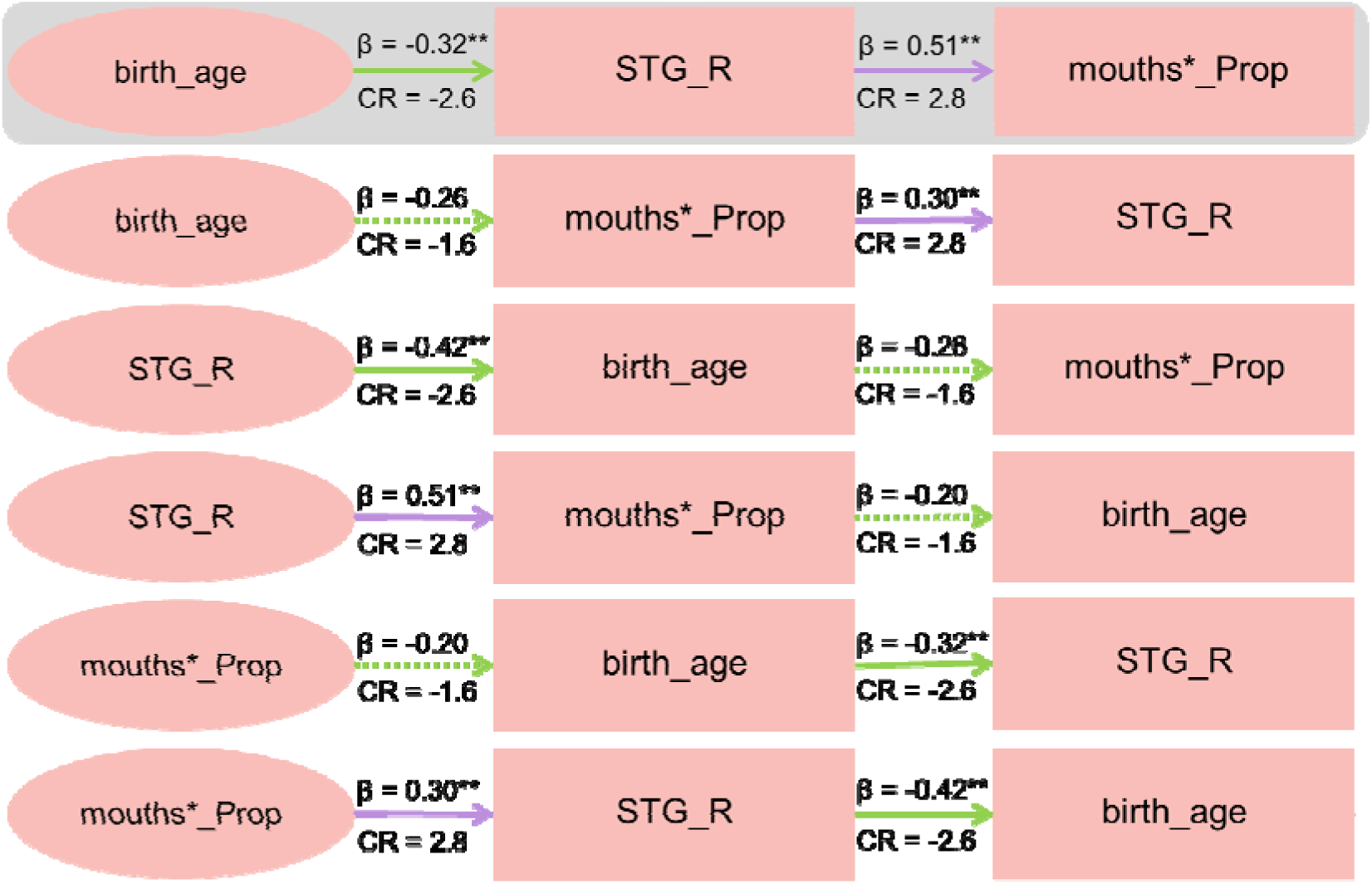
Path analysis in PB of age at birth, right STG GMV, and proportion of looking at mouths*. Solid line and dotted line each indicates a significant and non-significant path effect. Purple and green arrow each indicates positive and negative effect. The modeling included age at scan, TIV, sex, and the number of embryos at gestation as covariates. **p<0.01, CR: critical ratio. Note: For the sake of completeness, we evaluated all six models, although the models with proportion of looking at mouths* as independent variables were chronologically unlikely.

